# Prediabetes is associated with early-emerging and persistent cognitive differences in youth with overweight and obesity

**DOI:** 10.1101/2025.06.23.25330130

**Authors:** Jade Quillian, Filip Morys, Tuki Attuquayefio, Justin Sung, Antonietta Canna, Tiffany Ko, Xue Davis, Kaitlin Maciejewski, Fangyong Li, Nicola Santoro, Bridget Pierpont, Stephanie Kullmann, Hubert Preissl, Alain Dagher, Sonia Caprio, Dana M Small

## Abstract

Type 2 diabetes (T2D) is associated with cognitive impairment, but it remains unclear whether this reflects primary effects of metabolic dysfunction or secondary consequences of comorbid conditions, as most studies have focused on older adults. To address this, we examined the relationship between prediabetes and cognition in youth with overweight/obesity in a 2-year longitudinal study (n=67 at baseline; n=42 at follow-up). Participants underwent comprehensive cognitive testing, metabolic phenotyping, and structural and functional neuroimaging. At baseline, youth with prediabetes exhibited lower IQ and poorer performance in executive function, psychomotor speed, and visuospatial processing compared to those with normal glucose control, independent of adiposity. Across the full sample, reduced peripheral insulin sensitivity was associated with slower processing speed and altered central insulin sensitivity in the intraparietal sulcus. Longitudinally, cognitive differences between groups were stable over 2 years and were not explained by changes in adiposity or glucose tolerance. In contrast, expected developmental reductions in cortical surface area were observed only in youth with normal glucose control. These findings demonstrate that prediabetes in youth is associated with a broad and stable neurocognitive disadvantage that is independent of adiposity and not readily reversible with short-term changes in metabolic status. The early emergence and persistence of these differences suggest that altered neurocognitive function may be a feature of metabolic risk prior to overt disease onset.

## INTRODUCTION

Over 10% of the US and world adult populations have diabetes [1]. This number is estimated to increase to over 12% by 2045 with the majority (96%) of cases being individuals with type 2 diabetes (T2D) [2]. Since T2D increases the risk for developing dementia by 50% [3], these trends may contribute to the rapidly rising number of cases of Alzheimer’s disease (AD) and other dementias [4], [5]. Rates of T2D are also increasing in adolescents, with the prevalence doubling between 2001 and 2017 [6] and the age of onset continuing to shift to younger children [7]. Earlier ages of onset and increasing prevalence in adolescence provide a greater opportunity for the disease to negatively impact brain health and cognitive function in the population. This is due to both longer total disease duration and impact on the brain during a critical period of neurodevelopment. This is particularly concerning because cognitive ability is associated with academic achievement, which is in turn related to standard of living and a multitude of health outcomes [8].

Most of the work on neurocognition and metabolic dysfunction has focused on older adults. These studies report evidence of accelerated cognitive decline in those with T2D and prediabetes. This is true for general cognition [9], [10] as well as for domain specific functions. For example, T2D-associated cognitive decline is typified by impairment in executive function, psychomotor speed, attention, and memory [11], [12]. Prediabetes, in which individuals show impaired insulin sensitivity and less severe hyperglycemia, has also been associated with deficits in complex attention and executive function [13]. Since many of these functions are associated with dopaminergic signaling and alterations in the dopamine system are observed in obesity and T2D, we proposed that changes in the dopamine system may contribute to the association between T2D and cognition (Small 2017). Consistently, administration of intranasal insulin (INI) affects dopamine signaling in the striatum [14] and improves cognitive function in AD and mild cognitive impairment [15], [16], and points towards impaired central insulin sensitivity impacting dopamine signaling as a potential major contributing factor to cognitive decline in type 2 diabetes.

Structural neuroimaging studies in adults have also reported that T2D is associated with lower whole brain [17], [18], grey matter [19], [20], white matter [20], and hippocampal volume [19], [20], [21] as well as accelerated cortical atrophy and increases in ventricular volume [17], [18], [22]. Recent evidence suggests these differences exist in early adulthood and worsen with disease duration [23]. While this body of work provides unequivocal evidence for associations between T2D and brain health in adults, it is unclear if similar effects would be observed in younger individuals who are less likely to have comorbid conditions like cardiovascular disease that also impact brain health. The impact of diet and adiposity are also not well-considered [24], [25], [26].

In youth, there are few studies probing the associations between glucose metabolism and neurocognitive health, and those that have been done report conflicting results in neuroanatomical outcomes. For example, one study reported reduced white but not grey matter [27], while another reported the opposite [28]. There is more consistent evidence for cognitive deficits in tasks of executive functioning, processing/psychomotor speed, and verbal memory [27], [29]. Evidence of global cognitive deficits are reported in these studies, but only with abbreviated measures of IQ [27] or indirect measures such as academic achievement [29].

Another limitation with the current literature is that most studies are cross-sectional. Therefore, it remains unclear whether cognitive and brain changes result from: 1) direct metabolic effects of poor glucose control and insulin resistance, 2) comorbid conditions that accumulate with aging and disease duration (e.g., cardiovascular disease, hypertension), 3) shared risk factors such as obesity, poor diet, and sedentary lifestyle that independently affect both metabolic and brain health, or 4) reverse causality, whereby poorer cognitive function increases risk for developing T2D. Without longitudinal designs that control for adiposity and lifestyle factors, these mechanisms cannot be disentangled.

To address these issues, we recruited participants from the Pathogenesis of Youth Onset Diabetes study (PYOD). The PYOD is a longitudinal cohort of youth with overweight and obesity who have varying levels of glucose tolerance and insulin sensitivity. The PYOD provides an exceptional opportunity to study cognitive impairment because participants are pre-diabetic and thus do not suffer from chronic conditions associated with T2D. Cognition, peripheral glucose control, adiposity, brain structure and function and central insulin sensitivity were assessed at baseline and 2-years later. We address the following outstanding questions: 1) After controlling for adiposity, is prediabetes in youth associated with poorer general and domain specific cognitive outcomes? 2) Are these associations related to central insulin sensitivity, specifically in the brain’s dopaminergic system, and/or brain structure? 3) Are impairments greater in dopamine-dependent versus dopamine independent cognitive tasks? 4) Do these associations change over time? Specifically, are changes in glucose control associated with changes in cognition or brain structure/function?

## Methods

### Overall study description

The current report focuses on data collected during a two-year observational study in youth with overweight/obesity recruited primarily from the PYOD study. Participants completed an initial intake session, a four-day baseline assessment, three hour-long “short assessments” at 6-, 12-, and 18-month timepoints, and a four-day follow-up assessment at 24 months (Fig. 1a). The baseline and 24-month follow-up assessments both consisted of an oral glucose tolerance test and body composition measurement day, a comprehensive neuropsychological testing day, and two days of MRI scanning. The MRI days consisted of structural abdominal scans on one day and structural cranial scans on the other day, with a resting state functional magnetic resonance imaging (fMRI) procedure involving INI or placebo (counterbalanced across the two days), followed by cognitive tasks, performed on both days (Fig. 1b). Short assessments consisted of a fasted blood draw, cognitive tasks, and body composition measurements.

**Figure 1.**
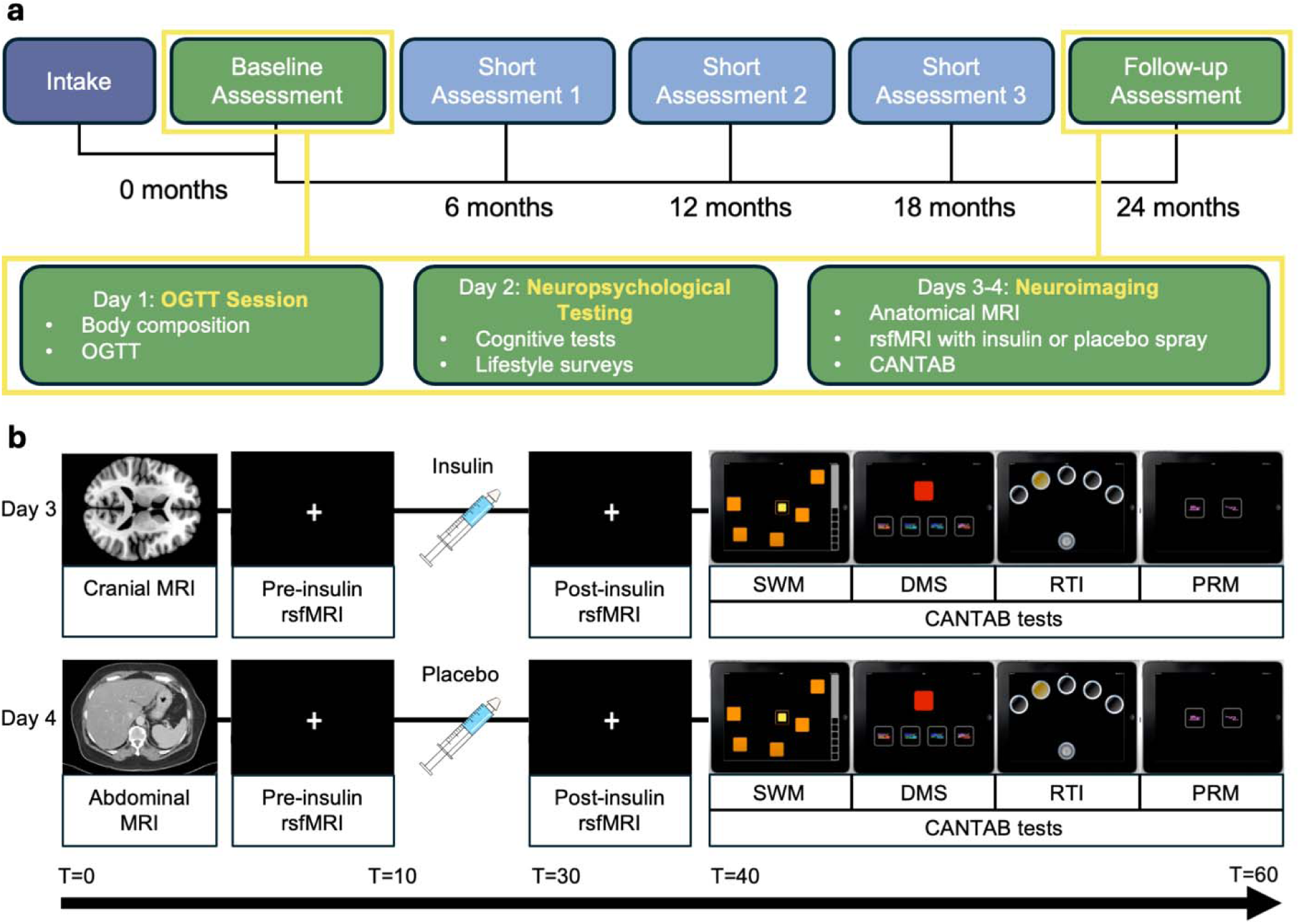
Study Design. (a) Overview of whole study timeline, highlighting the four-day Baseline Assessment and Follow-up Assessment that the present paper focuses on. (b) Resting state fMRI protocol with intranasal insulin. Order of administration of insulin and placebo was counterbalanced between days across participants.

### Participants

Adolescents with obesity (BMI above 85^th^ percentile) aged 8-20 were recruited from the ongoing PYOD study at Yale University and from physician referrals at Yale obesity outpatient clinics. Exclusion criteria included: diabetes, past head injury with loss of consciousness, developmental delays or cognitive deficits (not including ADHD), diagnosed psychiatric conditions, abnormal liver function, long-term or health history conditions that could impact study measures (i.e., cancer, hepatitis, etc.), and prior long-term medication usage that could impact study measures (i.e., metformin, SSRIs, etc.). Inclusion criteria were BMI > 85^th^ percentile and age 8-20 years.

All participants provided written informed consent/assent and were compensated. Those under 18 assented and parental consent was obtained. The study was conducted in accordance with the standards laid out in the Declaration of Helsinki. The study procedures were approved by the Yale Human Investigations Committee (Institutional Review Board Protocol no. 2000020794). The study was preregistered on 23 January 2018, to AsPredicted.org under access no. 7968, available at: https://aspredicted.org/ypx5-t6bp.pdf.

We had originally planned to complete 94 baseline assessments based on a power calculation for a 24-month analysis requiring 80 completers; however, enrollment and retention were negatively impacted by the COVID-19 pandemic. Sixty-nine participants completed all baseline assessment sessions and forty-three participants completed the follow-up assessment. A consort diagram depicting the number of participants from enrollment until baseline completion and analysis is presented in Supplement Figure 1.

### Study procedures

#### Intake session

After consent/assent was obtained from participants and parents when appropriate, participants underwent a mock MRI scan to familiarize them with the scanning environment, completed the Cambridge Automated Neuropsychological Test Battery (CANTAB; Cambridge Cognition Ltd) suite of tasks used in the study to assess executive function, psychomotor speed, and memory, and were administered an intranasal spray of saline to simulate the intranasal administration of insulin used in the study. Demographic data was collected at this session from participants or their parents.

Day 1: Oral glucose tolerance test (OGTT) session

Participants arrived after an overnight fast. First, body composition measurements were taken. Participants wore light clothing and removed shoes and socks for body composition measurements. Waist and hip circumference were measured manually by experimenters.

Height was measured by having participants stand straight with their back to a Seca stadiometer, which was lowered by experimenters until it touched the top of participant’s head. Weight, BMI, fat mass, and body fat percentage were measured using a connected Seca medical Body Composition Analyzer stand. Participants stood with feet making contact with electrodes on the base of the stand and lightly grabbed electrodes on the railing for bioelectric impedance analysis (BIA) used to determine fat mass and body fat percentage, while the machine also measured weight and calculated BMI. Age and sex adjusted BMI z-score and BMI 95^th^ percentile score were calculated according to CDC growth charts (https://www.cdc.gov/nchs/data/series/sr_11/sr11_246.pdf).

A 75g glucose OGTT was performed. An IV catheter was placed in the best vein found in the inner elbow of the arm or back of hand. A blood sample was collected to measure fasting glucose, insulin, and HbA_1C_ after which the oral glucose tolerance test was performed. Subjects were administered 75g of glucose in a drink consumed within 2 minutes. Subsequent blood samples were taken at 15, 30, 60, 90, and 120 minutes from which glucose and insulin concentrations were determined. Participants were classified as preT2D+ if they had two or three of the following impairments: impaired HbA_1C_ (>5.6), impaired fasting glucose (fasting glucose >100 mg/dl), and impaired glucose tolerance (2h glucose >140 mg/dl in the OGTT).

Otherwise, participants were classified as preT2D-. HOMA-IR was calculated using the following formula: (fasting glucose(mg/dL) * fasting insulin(µU/mL)/405. Matsuda index was calculated using insulin/glucose levels from five timepoints [30].

Day 2: neuropsychological assessment

Neuropsychological testing was conducted by trained staff in a quiet and distraction-free environment. Global and subdomain cognitive functioning was assessed using the Wechsler Intelligence Scale for Children, Fifth Edition (WISC-V) for participants under 16 years of age or the Wechsler Adult Intelligence Scale, Fourth Edition (WAIS-IV) for participants aged 16 years and older. Specifically, from this battery we analyzed composite cognition scores (Full Scale IQ, Fluid Reasoning Index, Verbal Comprehension Index) and sub-domain measures of executive functioning (Digit Span), psychomotor speed (Coding), and visuospatial processing (Block Design). Additionally, the Trail Making Test (TMT) A and B was administered to assess psychomotor speed (TMT A) and executive functioning (TMT B), an adolescent version of the Delayed Discounting Task (Wilson et al. 2010) was administered to assess temporal discounting, and the Variable Short Penn Line Orientation Test (VSPLOT; Gur et al. 2010) was administered to assess visuospatial perception. Further assessment of cognitive function was performed using the Cambridge Neuropsychological Test Automated Battery (CANTAB; Cambridge Cognition Ltd) administered using the Cambridge Connect application on an iPad. These tests were used to assess executive function (Spatial Working Memory, Stop Signal Task), psychomotor speed (Reaction Time Index), and memory (Pattern Recognition Memory, Delayed Matching to Sample). Diet was assessed via self-reported intake of fat and sugar in the Dietary Fat and Sugar Questionnaire (DFS; [31]). Surveys were also administered to assess depression (Center for Epidemiological Studies Depression Scale for Children, CES-DC [32]); and sleep apnea (Berlin Sleep Apnea Questionnaire, BSAQ [33]).

Due to ceiling effects on the original CANTAB spatial working memory task version used (4, 6, and 8-box trials), an extended 12 box version was added after the first 21 participants had completed this portion of the study at baseline. Only results from the 12-box version are used in this study’s analyses.

Days 3-4: scanner days - anatomical and functional neuroimaging

Participants arrived having fasted for at least two hours. Sessions began with anatomical images of the abdomen or head. Functional scans followed as depicted in Fig. 1b, with two 6-min resting-state functional magnetic resonance imaging (rsfMRI) scans, flanking (pre-/post spray scans) either INI (Humulin R human insulin 100 IU/ml, Lilly) or placebo plus a 15-min rest. Participants were taken out of the scanner for the INI/placebo spray. Insulin dose was of 1 IU per kg rounded to multiples of 10 IU (10, 20, 30 IU etc) up to a maximum of 60 IU, or an equivalent volume of sterile saline solution as placebo [34]. Insulin and placebo were administered using a syringe with an attached atomizer, spraying small puffs of solution into alternating nostrils while gently inhaling through the nostrils until the entirety of the solution had been inhaled. Immediately following the post-spray rsfMRI scan, participants were taken to a private testing room and completed our CANTAB test battery. The order in which participants received INI or placebo on the two scanner days was counterbalanced across subjects.

#### MRI acquisition

Anatomical and neuroimaging data were acquired on a Siemens Prisma 3 Tesla scanner at the Yale University Magnetic Resonance Research Center. Visceral and subcutaneous fat (deep and superficial) were assessed with a T1-weighted fast spin-echo sequence images of the abdomen. Proton Density Fat Fraction (PDFF) MRI liver images were acquired to assess liver fat fraction.

High resolution T1-weighted structural cranial images were acquired for each participant with repetition time (TR) = 1900 ms, echo time (TE) = 2.52 ms, flip angle = 9 1, field of view (FOV) = 250, matrix = 250 × 250, slice thickness = 1 mm, 176 slices, voxel size = 1× 1× 1 mm. To assess resting-state functional connectivity, whole-brain blood oxygenation level-dependent (BOLD) data were collected using multiband accelerated echo-planar imaging sequences, developed at Center for Magnetic Resonance Research (CMRR) Minnesota, USA. The functional images were acquired with TR = 1500 ms, TE = 34 ms, flip angle = 701, FOV = 192, partial Fourier=6/8, bandwidth= 1860 Hz/Px, echo spacing = 0.66 ms, matrix = 96 × 96, voxel size = 2× 2× 2 mm, images were acquired in interleaved order with a multiband acceleration factor of 4. Two other multi band scans with antero-posterior and poster-anterior phase encoding directions were acquired for distortion correction.

### Data analysis

Unless otherwise specified, statistical analysis was run using R studio (version 2024.12.1; R version 4.4.2). Outliers in the data were identified through visual inspection as well as values lying outside 2.2×interquartile range below the first quartile and above the third quartile.

Outlier values were assessed for experimental validity and analyses were repeated with these values excluded. No results changed significance based on inclusion or exclusion of outliers.

Final analyses presented are without outliers included.

### Missing data

At Baseline, Dietary Fat and Sugar surveys were missing responses on one item for six participants, and for two items for one subject, due to subjects skipping these questions. Scores for these missing data points were interpolated by calculating the mean response score for the subject within the relevant item category (sugar, fat, or sugar-and-fat). Otherwise, subjects with missing data for a given measure were excluded from analyses of that measure. Total household income was collected as a measure of socioeconomic status but was not included in our analyses because 18 subjects declined to respond. Group comparison of available data did not indicate differences in socioeconomic status (see Supplement Table 1 for data and comparison).

### Group characteristics

Sixty-nine subjects finished baseline assessments, with two subjects excluded from the analyses for having OGTT measurements (fasting glucose and 2h glucose) in the diabetic range. Of the sixty-seven remaining subjects, 44 were preT2D-, 22 were preT2D+, and one subject was unclassified due to missing OGTT data at baseline. These groups of 44 preT2D- and 22 preT2D+ were used for baseline group comparisons. At follow-up, one subject was excluded for having OGTT measurements in the diabetic range at baseline and one subject for missing OGTT data at baseline. Therefore, we had 42 usable datasets out of 43 completers, with a total of 27 preT2D-, 14 preT2D+, and one unclassified subject for our longitudinal comparisons. Group differences (preT2D+ vs preT2D-) were calculated using Welch Two Sample t-test, Pearson’s Chi-squared test, or Fisher’s exact test, as appropriate. Variables tested for baseline and follow-up group differences are listed in Table 1. Variables with a p < 0.10 at baseline were planned to be included as covariates in subsequent cross-sectional analyses, with IQ being the only one to reach this threshold (p=0.032).

**Table 1.** Characteristics of group, as defined at baseline. Comparison of demographic, adiposity, and metabolic measures between groups, as defined by prediabetes status at baseline, for both baseline and follow-up timepoints. *^1^* Welch Two Sample t-test, *^2^* Pearson’s Chi-squared test, *^3^* Fisher’s exact test.

|  | BASELINE CROSS-SECTIONAL |  |  |  | BASELINE LONGITUDINAL |  |  |  | FOLLOW-UP LONGITUDINAL |  |  |  |
| --- | --- | --- | --- | --- | --- | --- | --- | --- | --- | --- | --- | --- |
| Characteristic | Overall<br>N = 66 | preT2D-<br>N = 44 | preT2D+<br>N = 22 | p-value | Overall<br>N = 41 | preT2D-<br>N = 27 | preT2D+<br>N = 14 | p-value | Overall<br>N = 41 | preT2D-<br>N = 27 | preT2D+<br>N = 14 | p-value |
| Age |  |  |  | 0.87<sup>1</sup> |  |  |  | 0.6<sup>1</sup> |  |  |  | 0.6<sup>1</sup> |
| Mean (SD) | 14.24 (2.47) | 14.20 (2.37) | 14.32 (2.73) |  | 14.52 (2.59) | 14.36 (2.50) | 14.84 (2.84) |  | 16.65 (2.61) | 16.49 (2.51) | 16.96 (2.87) |  |
| Min, Max | 9.00, 20.00 | 9.00, 19.00 | 9.00, 20.00 |  | 9.66, 20.13 | 9.66, 18.07 | 9.69, 20.13 |  | 11.76, 22.18 | 11.84, 20.16 | 11.76, 22.18 |  |
| Sex |  |  |  | 0.60<sup>2</sup> |  |  |  | 0.9<sup>2</sup> |  |  |  |  |
| Female | 39 (59) | 25 (57) | 14 (64) |  | 24 (59%) | 16 (59%) | 8 (57%) |  |  |  |  |  |
| Male | 27 (41) | 19 (43) | 8 (36) |  | 17 (41%) | 11 (41%) | 6 (43%) |  |  |  |  |  |
| Race |  |  |  | 0.41<sup>3</sup> |  |  |  | 0.6<sup>3</sup> |  |  |  |  |
| Asian | 1 (1.5%) | 0 (0%) | 1 (4.55%) |  | 0 (0%) | 0 (0%) | 0 (0%) |  |  |  |  |  |
| Black/African-American | 20 (30.3%) | 11 (25%) | 9 (40.9%) |  | 11 (27%) | 6 (22%) | 5 (36%) |  |  |  |  |  |
| White | 26 (39.4%) | 19 (43.2%) | 7 (31.8%) |  | 19 (46%) | 12 (44%) | 7 (50%) |  |  |  |  |  |
| More than one race | 5 (7.6%) | 4 (9.1%) | 1 (4.55%) |  | 2 (4.9%) | 2 (7.4%) | 0 (0%) |  |  |  |  |  |
| Declined to respond | 14 (21.2%) | 10 (22.7%) | 4 (18.2%) |  | 9 (22%) | 7 (26%) | 2 (14%) |  |  |  |  |  |
| Ethnicity |  |  |  | >0.99<sup>2</sup> |  |  |  | 0.3<sup>3</sup> |  |  |  |  |
| Latino/Hispanic | 30 (48) | 20 (48) | 10 (48) |  | 18 (44%) | 10 (37%) | 8 (57%) |  |  |  |  |  |
| Non-Latino/Hispanic | 33 (52) | 22 (52) | 11 (52) |  | 21 (51%) | 16 (59%) | 5 (36%) |  |  |  |  |  |
| Decline to respond | 3 | 2 | 1 |  | 2 (4.9%) | 1 (3.7%) | 1 (7.1%) |  |  |  |  |  |
| BMI |  |  |  | 0.49<sup>1</sup> |  |  |  | >0.9<sup>1</sup> |  |  |  | 0.4<sup>1</sup> |
| Mean (SD) | 36 (7) | 37 (8) | 35 (6) |  | 36 (7) | 36 (8) | 36 (7) |  | 39 (8) | 40 (9) | 38 (7) |  |
| Min, Max | 23, 52 | 23, 52 | 25, 51 |  | 23, 52 | 23, 52 | 25, 51 |  | 27, 59 | 27, 59 | 28, 57 |  |
| BMI z-score |  |  |  | 0.94<sup>1</sup> |  |  |  | 0.8<sup>1</sup> |  |  |  | >0.9<sup>1</sup> |
| Mean (SD) | 2.29 (0.42) | 2.30 (0.44) | 2.29 (0.39) |  | 2.31 (0.42) | 2.30 (0.42) | 2.33 (0.44) |  | 2.37 (0.46) | 2.36 (0.48) | 2.38 (0.43) |  |
| Min, Max | 1.23, 2.95 | 1.52, 2.95 | 1.23, 2.81 |  | 1.23, 2.88 | 1.52, 2.88 | 1.23, 2.81 |  | 1.48, 3.21 | 1.48, 3.21 | 1.48, 3.05 |  |
| missing |  |  |  |  |  |  |  |  | 1 | 0 | 1 |  |
| BMI 95<sup>th</sup> Percentile |  |  |  | 0.55<sup>1</sup> |  |  |  | >0.9<sup>1</sup> |  |  |  | 0.6<sup>1</sup> |
| Mean (SD) | 133 (25) | 134 (27) | 131 (21) |  | 135 (25) | 135 (26) | 135 (23) |  | 139 (28) | 140 (30) | 136 (25) |  |
| Min, Max | 89, 190 | 96, 190 | 89, 177 |  | 89, 190 | 96, 190 | 89, 177 |  | 95, 202 | 95, 202 | 95, 188 |  |
| missing |  |  |  |  |  |  |  |  | 1 | 0 | 1 |  |
| Waist to Hip Ratio |  |  |  | 0.28<sup>1</sup> |  |  |  | 0.3<sup>1</sup> |  |  |  | 0.15<sup>1</sup> |
| Mean (SD) | 0.88 (0.06) | 0.87 (0.06) | 0.89 (0.05) |  | 0.88 (0.05) | 0.87 (0.06) | 0.89 (0.04) |  | 0.91 (0.05) | 0.90 (0.05) | 0.92 (0.05) |  |
| Min, Max | 0.79, 1.02 | 0.79, 1.02 | 0.79, 0.98 |  | 0.79, 1.02 | 0.79, 1.02 | 0.81, 0.95 |  | 0.83, 1.01 | 0.83, 1.01 | 0.85, 1.01 |  |
| missing | 3 | 3 | 0 |  | 2 | 2 | 0 |  |  |  |  |  |
| Fat Mass (kg) |  |  |  | 0.39<sup>1</sup> |  |  |  | 0.8<sup>1</sup> |  |  |  | 0.4<sup>1</sup> |
| Mean (SD) | 41 (14) | 42 (16) | 39 (12) |  | 40 (15) | 41 (15) | 39 (14) |  | 51 (19) | 52 (21) | 47 (14) |  |
| Min, Max | 11, 72 | 11, 72 | 19, 65 |  | 11, 69 | 11, 69 | 19, 65 |  | 21, 108 | 21, 108 | 25, 76 |  |
| missing | 6 | 4 | 2 |  | 5 | 3 | 2 |  |  |  |  |  |
| Fat percentage (%) |  |  |  | 0.81<sup>1</sup> |  |  |  | 0.7<sup>1</sup> |  |  |  | 0.7<sup>1</sup> |
| Mean (SD) | 43 (7) | 43 (8) | 43 (6) |  | 42 (8) | 42 (8) | 41 (7) |  | 45 (8) | 45 (8) | 46 (6) |  |
| Min, Max | 24, 61 | 24, 61 | 30, 53 |  | 24, 61 | 24, 61 | 30, 53 |  | 29, 60 | 29, 60 | 34, 57 |  |
| missing | 6 | 4 | 2 |  | 7 | 3 | 4 |  |  |  |  |  |
| Visceral to subcutaneous fat ratio |  |  |  | 0.12<sup>†</sup> |  |  |  | 0.3<sup>†</sup> |  |  |  | 0.3<sup>†</sup> |
| Mean (SD) | 0.17 (0.06) | 0.16 (0.06) | 0.19 (0.06) |  | 0.16 (0.05) | 0.16 (0.06) | 0.17 (0.04) |  | 0.18 (0.05) | 0.19 (0.05) | 0.16 (0.05) |  |
| Min, Max | 0.07, 0.36 | 0.07, 0.36 | 0.09, 0.30 |  | 0.07, 0.36 | 0.07, 0.36 | 0.10, 0.22 |  | 0.11, 0.31 | 0.11, 0.31 | 0.11, 0.24 |  |
| missing | 4 | 4 | 0 |  | 2 | 2 | 0 |  | 9 | 5 | 4 |  |
| Deep to superficial subcutaneous fat ratio |  |  |  | 0.10<sup>†</sup> |  |  |  | 0.054<sup>†</sup> |  |  |  | 0.018<sup>†</sup> |
| Mean (SD) | 1.47 (0.53) | 1.55 (0.57) | 1.33 (0.44) |  | 1.54 (0.57) | 1.66 (0.61) | 1.33 (0.44) |  | 1.65 (0.48) | 1.76 (0.51) | 1.41 (0.29) |  |
| Min, Max | 0.71, 3.88 | 0.75, 3.88 | 0.71, 2.40 |  | 0.71, 3.88 | 0.88, 3.88 | 0.71, 2.40 |  | 0.94, 3.03 | 0.95, 3.03 | 0.94, 1.88 |  |
| missing | 4 | 4 | 0 |  | 2 | 2 | 0 |  | 9 | 5 | 4 |  |
| Liver fat fraction |  |  |  | 0.15<sup>†</sup> |  |  |  | 0.8<sup>†</sup> |  |  |  | 0.3<sup>†</sup> |
| Mean (SD) | 5.3 (6.0) | 4.4 (4.8) | 6.9 (7.5) |  | 5.1 (5.6) | 4.9 (5.8) | 5.4 (5.4) |  | 7 (8) | 7 (9) | 5 (5) |  |
| Min, Max | 0.1, 31.0 | 0.1, 21.1 | 0.9, 31.0 |  | 0.1, 21.1 | 0.1, 21.1 | 1.4, 19.9 |  | 1, 30 | 1, 30 | 1, 18 |  |
| missing | 3 | 3 | 0 |  | 5 | 3 | 2 |  | 6 | 4 | 2 |  |
| DFS survey total |  |  |  | 0.11<sup>†</sup> |  |  |  | 0.2<sup>†</sup> |  |  |  | 0.9<sup>†</sup> |
| Mean (SD) | 55 (13) | 57 (13) | 52 (12) |  | 52 (11) | 53 (12) | 49 (10) |  | 56 (11) | 56 (12) | 55 (10) |  |
| Min, Max | 28, 82 | 28, 82 | 36, 81 |  | 28, 77 | 28, 77 | 36, 67 |  | 34, 84 | 34, 84 | 41, 77 |  |
| missing | 3 | 1 | 2 |  | 2 | 0 | 2 |  |  |  |  |  |
| Matsuda index |  |  |  | 0.002<sup>†</sup> |  |  |  | 0.049<sup>†</sup> |  |  |  | 0.8<sup>†</sup> |
| Mean (SD) | 1.44 (0.86) | 1.64 (0.97) | 1.07 (0.42) |  | 1.53 (0.99) | 1.71 (1.15) | 1.19 (0.46) |  | 1.72 (2.23) | 1.76 (2.64) | 1.63 (1.08) |  |
| Min, Max | 0.56, 5.40 | 0.69, 5.40 | 0.56, 1.96 |  | 0.63, 5.40 | 0.69, 5.40 | 0.63, 1.96 |  | 0.40, 14.23 | 0.40, 14.23 | 0.42, 4.62 |  |
| Missing | 1 | 1 | 0 |  |  |  |  |  | 2 | 1 | 1 |  |
| HbA1c |  |  |  | 0.46<sup>†</sup> |  |  |  | >0.9<sup>†</sup> |  |  |  | 0.6<sup>†</sup> |
| Mean (SD) | 5.28 (0.45) | 5.26 (0.47) | 5.34 (0.42) |  | 5.28 (0.43) | 5.29 (0.42) | 5.28 (0.48) |  | 5.32 (0.47) | 5.36 (0.42) | 5.26 (0.58) |  |
| Min, Max | 4.00, 6.20 | 4.00, 6.20 | 4.60, 6.00 |  | 4.60, 6.20 | 4.60, 6.20 | 4.60, 6.00 |  | 4.50, 6.70 | 4.80, 6.50 | 4.50, 6.70 |  |
| Missing | 1 | 1 | 0 |  |  |  |  |  | 2 | 2 | 0 |  |
| Fasting Glucose (mg/dl) |  |  |  | <0.001<sup>†</sup> |  |  |  | 0.003<sup>†</sup> |  |  |  | 0.3<sup>†</sup> |
| Mean (SD) | 102 (10) | 99 (9) | 109 (8) |  | 100 (9) | 98 (9) | 105 (5) |  | 103 (12) | 105 (11) | 100 (13) |  |
| Min, Max | 80, 123 | 80, 122 | 96, 123 |  | 80, 122 | 80, 122 | 96, 116 |  | 77, 127 | 87, 127 | 77, 126 |  |
| Missing |  |  |  |  |  |  |  |  | 1 | 1 | 0 |  |
| OGTT 2h Glucose |  |  |  | <0.001<sup>†</sup> |  |  |  | <0.001<sup>†</sup> |  |  |  | 0.8<sup>†</sup> |
| Mean (SD) | 131 (23) | 119 (14) | 155 (19) |  | 131 (26) | 116 (15) | 158 (20) |  | 135 (34) | 134 (34) | 137 (35) |  |
| Min, Max | 78, 196 | 78, 137 | 122, 196 |  | 78, 196 | 78, 137 | 123, 196 |  | 56, 201 | 56, 197 | 77, 201 |  |
| Missing | 1 | 1 | 0 |  |  |  |  |  | 2 | 2 | 0 |  |
| Full Scale IQ |  |  |  | 0.032<sup>†</sup> |  |  |  | 0.016<sup>†</sup> |  |  |  | 0.016<sup>†</sup> |
| Mean (SD) | 97 (14) | 99 (15) | 92 (10) |  | 99 (15) | 102 (16) | 92 (10) |  | 98 (16) | 102 (16) | 91 (12) |  |
| Min, Max | 70, 132 | 70, 132 | 78, 110 |  | 70, 132 | 70, 132 | 78, 108 |  | 68, 139 | 71, 139 | 68, 109 |  |

### Association between adiposity and metabolic measures

To determine the associations between adiposity and metabolic measures, we ran Pearson correlations across measures and created a correlation matrix using GraphPad Prism 9 software (GraphPad Software 2021).

### Data visualization

Graphs were made using R studio (version 2024.12.1). MRI visualizations were made using MRIcroGL (obtained from https://www.nitrc.org/projects/mricrogl/).

### Cross-sectional cognition analyses

Group differences in cognition across domains:

Linear regressions were run to assess group differences in performance on tasks of various cognitive domains, with age, sex, and BMI z-score as covariates (model 1) and with IQ added as an additional covariate (model 2) so that we could identify deficits relative to IQ. Benjamini-Hochberg correction (BHC) was applied to domains with multiple measures to account for multiple comparisons (α=0.05 for this and all following analyses).

### Scanner days: effects of intranasal insulin (INI) administration

The effect of insulin on cognitive performance within and between groups was assessed for the CANTAB battery (stop signal task not included in this testing for time). Linear mixed model analyses were used to investigate the effect of group (preT2D-/preT2D+), condition (INI/Placebo) and group by condition interactions. The models included participant ID as a random effect to account for the repeated measurements done on each participant (insulin vs placebo).

### Effects of peripheral insulin sensitivity on cognition (exploratory analyses)

We performed exploratory linear regressions on Matsuda index’s relation to cognition across the full sample with age and BMI z-score as covariates. We also performed linear mixed model analyses to assess the effects of interactions between conditions (insulin vs placebo) and Matsuda index on performance on the CANTAB battery assessed on the imaging days. Age, sex, and BMI z-score were used as covariates; participant ID was used as a random effect.

### Longitudinal analysis of cognition

Change in prediabetes status and cognitive variability over time:

To test for effects related to change in prediabetes status we classified subjects at follow-up as preT2D- and preT2D+ using the same criterion as with the baseline data. Some participants improved in their metabolic health over the two years (i.e., preT2D+ classification at baseline to preT2D- classification at follow-up) while others worsened (i.e., preT2D- to preT2D+). We hypothesized that change in metabolic status would be accompanied by a greater magnitude of variability in cognition. We therefore compared the absolute value of the difference between cognitive task scores from the Neuropsychological Assessment at follow-up and baseline for those who changed group classification versus those who did not. We used absolute values to account for the fact that individuals whose metabolic health improved might have changed their cognition in a different direction than individuals whose metabolic health worsened. We combined those participants (whose metabolic health improved and worsened) into one group due to low sample size of separate groups. Linear regression models tested the effect of group (changed vs stable metabolic health) on magnitude of baseline vs follow-up differences (absolute values) in cognitive performance with age at baseline, BMI z-score at baseline, sex, and task score at baseline as covariates.

Effects of baseline prediabetes status, change in insulin resistance, and change in adiposity on cognition over time:

We next tested for effects of baseline group assignment (T2D+ vs. T2D-) on measures of cognition over time to examine if prediabetes is predictive of cognitive change, irrespective of change in metabolic health. Linear mixed effects models were run to assess effects of baseline group classification on cognitive performance (main effect), as well as change in cognition over time (group X time interaction). Separate analogical analyses were performed using adiposity and insulin resistance measures instead of group classification. For all models, age and sex were used as fixed effects and participant ID was included as a random effect to account for repeated measures. For the analysis looking at group, BMI was included as a covariate to control for effects of adiposity. Multiple comparisons correction was performed using BHC on p-values across tasks within the same cognitive domain grouping (see Supplement Table 2 for groupings) for main effects or main effect X time interactions within each group, Matsuda, or BMI set of models.

### Cross-sectional resting state fMRI correlates

MRI data preprocessing

Functional data were preprocessed using DPARSF v. 5.4 (https://rfmri.org/DPARSF), SPM12 (https://www.fil.ion.ucl.ac.uk/spm/software/spm12/) running on MATLAB 2016a and FSL (https://fsl.fmrib.ox.ac.uk/fsl/fslwiki/FslInstallation). Sample size was limited by subject exclusions due to excessive motion in scanner and available data for main effects. Supplement Table 3 summarizes sample sizes for analyses run on rsfMRI data.

### Preprocessing

By evaluating the motion parameters, only those datasets surviving the exclusion-criteria of a threshold of 4mm for translation and 4 degrees for rotation, plus a threshold on the framewise displacement measurement to be less than 0.5 were considered for the other (pre)-processing steps and statistical analyses. Data for participants in which at least one of the four sessions did not survive the movement criteria were discarded for analyses involving time (pre- vs post-spray) as a factor. Data for participants in which at least one of the two post-spray sessions did not survive the movement criteria were discarded for all analyses.

The considered data were then corrected for field distortion using the TOPUP tool of FSL [35]. All data from all 4 sessions were aligned across each other (using the co-registration implemented in SPM), and then nuisance [36], [37], [38] and Friston24 regressions [39] as well as motion scrubbing [39], [40] were applied using the fsl_glm algorithm of FSL. For the nuisance regression, white matter (WM) and cerebrospinal fluid (CSF) masks were obtained from the T1 segmentation (as part of the DPARFS pipeline). These two masks were 90% thresholded, binarized and aligned to the functional data (using the co-registration implemented in SPM). Mean values of the functional data within all voxels of these masks were then computed. Data were normalized in the MNI standard space using the normalize algorithm implemented in SPM and resampled to 2mm isotropic voxels. In the standard space, temporal filtering with a band pass filter with cut off frequencies between 0.01 and 0.1 Hz, and spatial smoothing with a gaussian kernel with a 6mm size, were applied.

### Fractional amplitude of low frequency fluctuations (fALFF) analyses

To assess the impact of INI of resting state brain activity at baseline, fractional amplitude of low frequency fluctuations [41] was implemented in the DPARFS toolbox. The relative intensity of BOLD fluctuations at rest in a given area was determined by the ratio of the power spectrum of the defined low-frequency range (0.01-0.1Hz) taken in relation to the power of the entire frequency range (0-0.25Hz). As previously suggested [42], data without temporal filtering were used to compute the fALFF maps.

Statistical analyses were performed in SPM12. We had originally planned to use a full factorial design that included time (2 levels, pre and post intranasal injection), condition (2 levels: placebo and INI), and group (2 levels: preT2D+ and preT2D-) as within subjects’ factors and age, sex, and BMI z-score as covariates for the subgroup of participants where all four scans passed our criterion for acceptable movement. However, only 19 T2D- and 9 preT2D+ participants passed this criterion. Therefore, we abandoned this approach and limited further analyses to regressions with post-spray scans only and collapsed across groups, resulting in a larger sample size. First, the difference in fALFF values between insulin and placebo conditions was calculated for each subject in SPM using a voxel-based subtraction of the post-insulin scan minus the post-placebo scan. The resulting insulin-placebo maps were used in a multiple regression analysis with Matsuda index (n=41), using age, sex, and BMI z-score as covariates. Using the same covariates, multiple regression analyses were run on scores from tasks conducted during the neuropsychological testing session where significant differences were observed between groups; spatial working memory (n=28), reaction time index (n=40), block design (n=42), VSPLOT (n=37), and stop signal task (n=38). Sample sizes differed due to missing or invalid data. Finally, the difference in the scores from the four tests completed on rsfMRI scan days (spatial working memory, delayed match-to-sample, pattern recognition memory, and reaction time index) were calculated for post-insulin minus post-placebo, and multiple regression analyses were run, again using age, sex, and BMI z-score as covariates. Contrast estimates were extracted from the peak voxel of every significant cluster for each of these analyses from the insulin-placebo maps to be plotted against their relevant variables for visualization of the direction of associations.

### Seed-based caudate connectivity analyses

To assess the impact of INI on dopamine-dependent signaling and cognitive function, seed-based analyses of caudate connectivity were employed. For each participant, seed-based correlation maps were computed using the Pearson correlation between the signal time course of each voxel and the mean signal time course in the seed, located in the bilateral caudate and obtained using the wfu_pickatlas tool (https://www.nitrc.org/projects/wfu_pickatlas). Finally, the correlation values were transformed to z-values using the Fisher transformation.

In the same manner as above with the fALFF analysis, insulin-placebo maps were calculated for each subject and regression analyses were run to determine effects of condition and group on caudate connectivity under intranasal insulin. Regressions were run for scores from the three tasks from the neuropsychological test day that showed significant differences and for all four tests completed on the rsfMRI scan days.

### Brain structure analyses

MRI data preprocessing

Structural T1-weighted images were corrected for gradient non-linearity distortions and processed with FreeSurfer v. 7.4.1 (https://surfer.nmr.mgh.harvard.edu/) using the -recon_all function and FreeSurfer’s longitudinal pipeline. We derived global measures of cortical thickness and surface area as described below. Additionally, we used the Destrieux parcellation data [43] to obtain information on cortical thickness of the intraparietal sulcus (IPS), as well as Freesurfer’s automatic subcortical segmentation to obtain the volume of the hippocampus.

These regions of interest were based on previous reports of reduced hippocampal volume in individuals with type 2 diabetes [19], [20], [21], [23] and findings in this manuscript. Cortical and subcortical segmentation was visually inspected for quality control using pass/fail ratings, where those that did not pass were excluded from analysis.

Anatomical measures of interest were calculated as follows:

Global mean cortical thickness

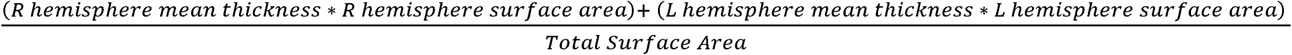

Total surface area

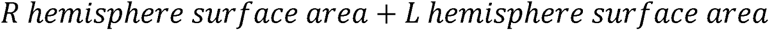

IPS thickness

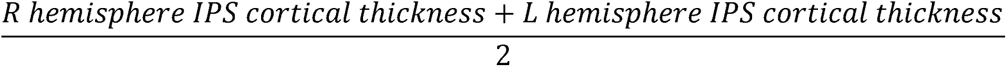

Hippocampal volume

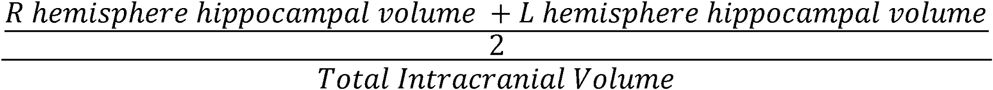

Cross-sectional analysis

Of the sixty-seven subjects included in baseline analyses, seven subjects were excluded from cortical anatomical analyses at baseline due to MRI artifacts. Of the 60 remaining anatomical datasets, 39 were preT2D-, 20 were preT2D+, and one subject was unclassified due to missing OGTT data at baseline. These groups of 39 preT2D- and 20 preT2D+ were used for baseline cortical anatomical comparisons.

Separate linear regression models were used to assess effects of baseline prediabetes group (preT2D+ vs. pre T2D-), baseline insulin resistance (Matsuda index), and baseline adiposity (BMI z-score) on our primary anatomical measures of interest (global mean cortical thickness, total surface area, IPS thickness, and hippocampal volume). Age and sex were used as covariates for all models, with BMI z-score added as a covariate for the group analysis to control for adiposity. Multiple comparisons correction was performed using BHC on main effect p-values across group, BMI z-score, and Matsuda models for each anatomical measure.

### Longitudinal analysis

Twelve participants were not scanned at follow-up due to newly acquired contraindications such as dental braces or not being able to fit in the scanner. Four follow-up completers were additionally excluded at baseline from anatomical analysis due to artifacts as previously mentioned. Therefore 26 of 42 subjects had analyzable anatomical data for longitudinal analysis, with a total of 15 preT2D-, 10 preT2D+, 1 without group classification.

Separate linear mixed model regressions were similarly used to assess effects of the baseline prediabetes group (preT2D+ vs. preT2D-), insulin resistance (Matsuda index), and adiposity (BMI) on the aforementioned four brain region measures. Similar to our analysis on longitudinal cognition, we used baseline prediabetes status to determine the predictive role of prediabetes classification, irrespective of change in metabolic status. Furthermore, we investigated changes in brain structure over time related to changes in Matsuda index and BMI across both timepoints to assess the longitudinal effects of insulin resistance and adiposity. Time, age, and sex were used as fixed effect covariates and participant ID was used as a random effect for all models, with BMI added as a covariate for the group analysis to control for adiposity. Multiple comparisons correction was performed using BHC on main effect p-values and main effect by time interaction p-values across group, BMI z-score, and Matsuda models for each anatomical measure.

## RESULTS

### Cross-sectional results (Group characteristics and Behavioral)

#### Cross-sectional characteristics of groups at baseline

PreT2D- (n=44) and preT2D+, (n=22) groups did not differ on any of the demographic, dietary, adiposity, sleep, or depression measures (p≥0.1). Mean Full Scale IQ differed significantly between groups (Fig. 2a, t(57.6) = 2.2, mean difference = 7 95% CI [0.64, 13], p = 0.032), with the preT2D+ group (mean=92, SD=10.41) scoring lower than the preT2D- group (mean = 99 ± 15.16). Insulin sensitivity, as measured by Matsuda index, in preT2D+ (mean = 1.07± 0.42) was significantly lower (Fig. 2b, t(62) = 3.31, mean difference = 0.57 95% CI [0.23, 0.92], p = 0.002) than in preT2D- (mean = 1.67 ± 0.97). Full results reported in Table 1.

**Figure 2.**
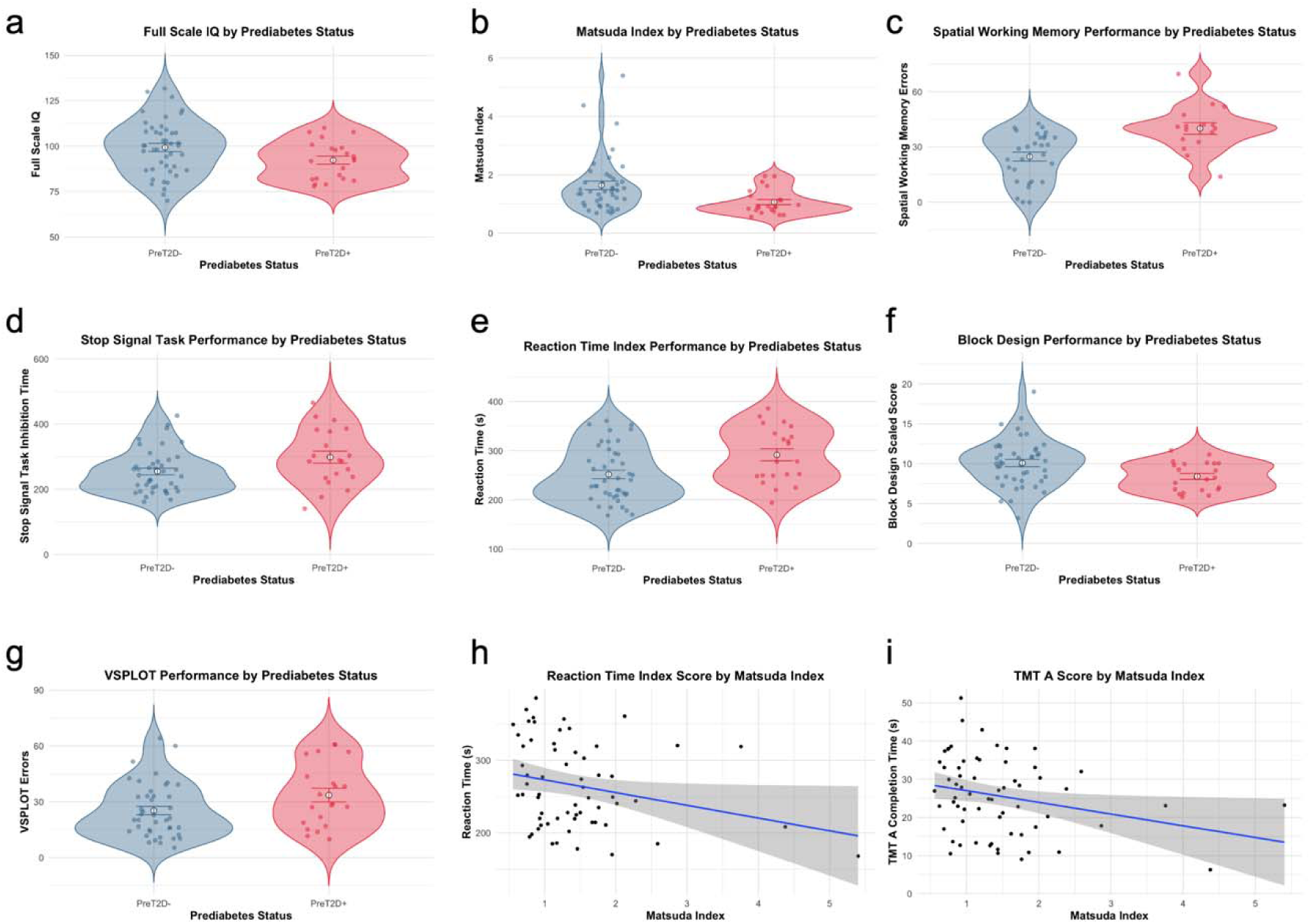
Baseline differences between groups. Participants with prediabetes had significantly lower Full Scale IQ (a) and insulin sensitivity as measured by Matsuda Index (b). Participants with prediabetes performed worse on (c) the CANTAB Spatial Working Memory Task (version with 12 boxes), (d) the CANTAB Stop Signal Task, (e) CANTAB Reaction Time Index task, (f) the WISC/WAIS Block Design (BD) task, and (g) the Variable Short Penn Line Orientation Task (VSPLOT). Matsuda index is negatively correlated with (h) Reaction Time Index and (i) Trail Making Test A (task performance.

#### Association between adiposity and metabolic measures

To identify associations between adiposity and metabolic measures, we used Pearson correlation analysis. We found that Matsuda Index correlated with almost all measures of both glucose metabolism and adiposity (Supplement Figure 2). Conversely, HbA1c did not correlate with other measures. HOMA-IR correlated with both BMI-z score and with measures of abdominal adiposity. Adiposity measures were highly correlated within themselves.

#### Group differences in cognition across domains

To test the prediction that metabolic health is related to cognition independent of adiposity, we ran linear regression analyses with group as the predictor of interest, cognitive tasks from the neuropsychological assessment (Supplement Table 2) as outcome variables, and age, sex, and BMI z-scores as covariates. Consistent with our prediction, our analysis revealed impaired executive functioning, psychomotor speed, and visuospatial processing in preT2D+ compared to preT2D-(Fig. 2c-g). Specifically, compared to preT2D-, preT2D+ had: 1) more errors on the spatial working memory task (Fig. 2c, preT2D+ mean = 40.00 ± 12.61, preT2D- mean = 24.66 ± 13.21, estimate = 14.753 ± 4.036, p < 0.001); 2) a larger 50% reaction inhibition time was larger on the stop signal task, indicating greater impulsivity (Fig. 2d, preT2D+ mean = 298.01 ± 85.71, preT2D- mean = 254.36 ± 65.79, estimate = 44.300 ± 18.133, p = 0.0177); 3) a slower reaction time index (Fig. 2e, preT2D+ mean = 291.57 ± 56.49, preT2D- mean = 251.81 ± 56.05, estimate = 39.909 ± 14.903, p = 0.00962); 4) a lower score on block design (Fig. 2f, preT2D+ mean = 8.41 ± 1.79, preT2D- mean= 10.09 ± 3.05, estimate = −1.672 ± 0.723, p = 0.0241); and 5) poorer performance on the variable short Penn line orientation test, indicating poorer early visual spatial processing (Fig. 2g, preT2D+ mean = 33.62 ± 17.03, preT2D- mean= 25.34 ± 14.57, estimate = 8.647 ± 3.843, p = 0.0283). We also re-ran these analyses after including IQ as an additional covariate in the regressions to identify if there were cognitive effects that occur independently of the decreased IQ in preT2D+. Only spatial working memory survived this analysis (estimate = 12.734 ± 4.131, p = 0.00376), suggesting that the primary effect is a global decrease in cognition, with the exception of spatial working memory.

#### Scanner day: effects of intranasal insulin (INI) administration on cognition

To test if INI administration affects cognition and to understand whether the effects depend on metabolic health, we used linear mixed model analyses with group (preT2D-/preT2D+), condition (INI/Placebo), and a group by condition interaction as regressors of interest. We observed a significant effect of group on reaction time index performance (F(1,64)=3.89, p = 0.047), but no significant effects of condition on performance or two-way interactions between group and condition on task performance. This indicates that, contrary to our prediction, INI treatment did not impact performance, apart from processing speed, on the cognitive tasks that we administered during the imaging sessions.

#### Effects of peripheral insulin sensitivity on cognition (exploratory analyses)

To test if peripheral insulin sensitivity is associated with cognition across the full sample (preT2D+ and preT2D-), we ran linear regression analyses using Matsuda index as predictor and cognitive measures assessed during the neuropsychology day as outcome variables with age, sex, and BMI z-scores as covariates. The only significant effects to emerge were negative associations between Matsuda index and scores on reaction time index (Fig. 2h, β = −20 95% CI [−38, −0.58], p = 0.044) and Trail Making Test A (Fig. 2i, β = −4.12 95% CI [−6.99, −1.26], p = 0.005471), consistent with a robust effect of peripheral glucose control on processing speed.

We also tested if peripheral insulin sensitivity might influence the ability of INI to influence cognition across the whole group. We used linear mixed effects models with condition (INI/Placebo) as an additional predictor and participant ID as a random effect. No significant effects were observed between peripheral insulin sensitivity (Matsuda index) and cognitive performance under INI vs placebo.

### Cross-sectional results (neuroimaging)

#### fALFF-peripheral metabolic health and central insulin sensitivity

To test for associations between metabolic health and central insulin sensitivity, Matsuda index was regressed against resting state brain activity (fALFF) in insulin minus placebo. Age, sex, and BMI z-score served as covariates for all rsfMRI analyses. This identified a positive correlation between peripheral insulin sensitivity and central insulin sensitivity in the left intraparietal sulcus (MNI coordinates: −26, −52, 40; k=54; p_(FWE)_ =0.048) (Fig. 3a-b).

**Figure 3.**
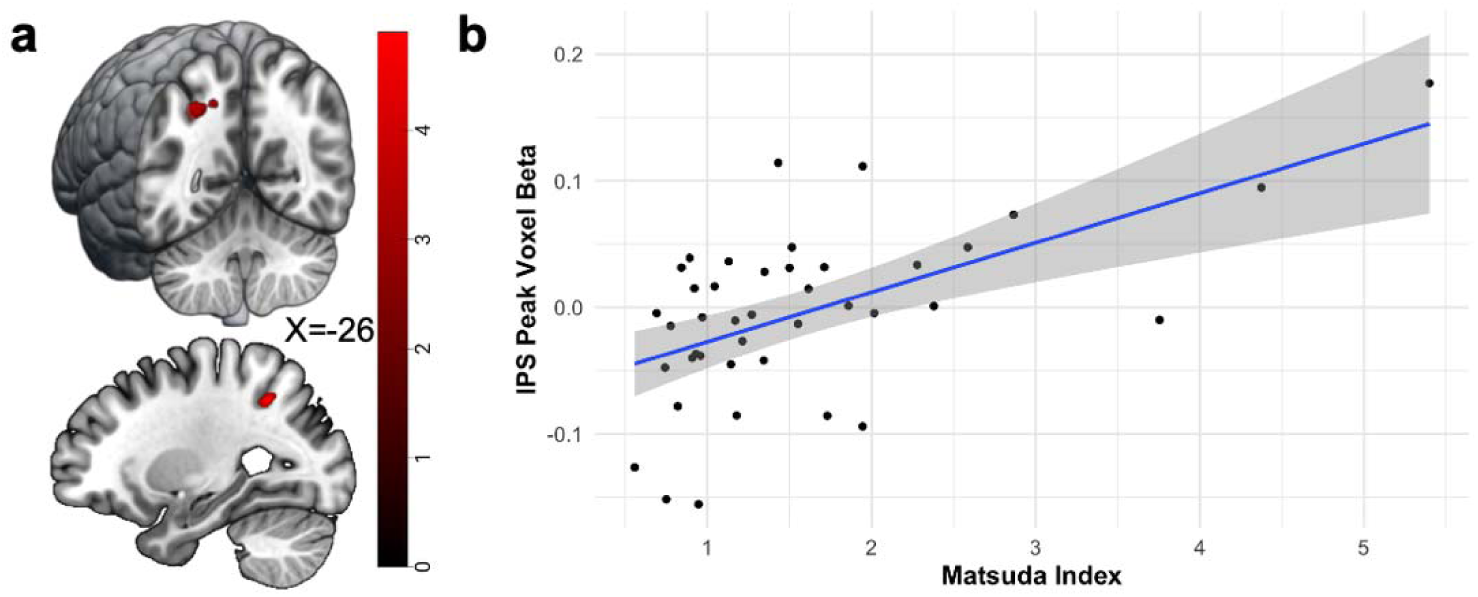
Peripheral metabolism effects on resting state brain activity. (a) Correlation between Matsuda Index and response to INI in the left intraparietal sulcus (MNI coordinates: −26 −52 40). (b) Plotted values of the peak voxel from the cluster in a from the Insulin – Placebo map against Matsuda index.

#### fALFF- Effects related to cognition

As outlined above, poor peripheral glucose control was associated with poorer performance in spatial working memory, reaction time index, Penn line orientation and block design tasks. To identify potential neural correlates for these cognitive effects we regressed performance with resting state activation under insulin vs placebo administration. The only effect to emerge was within the right medial prefrontal cortex (MNI coordinates: 12, 56, 2; k=61; p_(FWE)_ =0.032), which was inversely associated with reaction time from the reaction time index (Fig. 4a and b), indicates that insulin sensitivity in the right medial prefrontal cortex is associated with the reduced processing speed.

**Figure 4.**
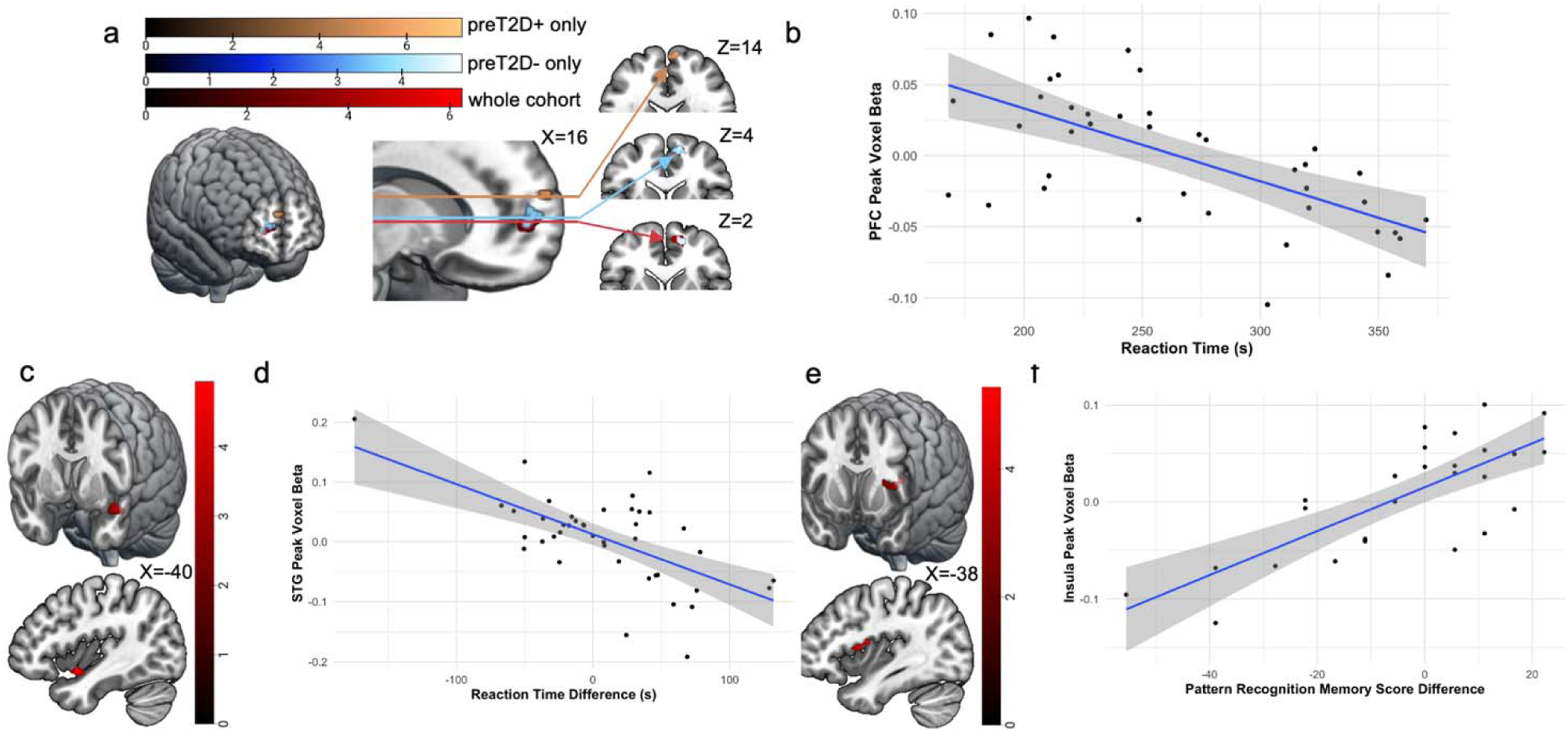
Brain sensitivity to INI related to cognitive performance. (a) Right medial anterior prefrontal cortex (MNI coordinates: 12 56 2) INI sensitivity is negatively correlated with Reaction Time Index performance in whole cohort. Non-significant anterior medial prefrontal cortex cluster (MNI coordinates: 8 64 14) in regression of PD group RTI performance and INI sensitivity. (MNI coordinates: 12 58 4) Significant anterior medial prefrontal cortex cluster (MNI coordinates: 8 64 14) in regression of NPD group RTI performance and INI sensitivity. (b) Plotted values of the peak voxel from the whole cohort cluster in a. (c) Left supratemporal gyrus (MNI coordinates: −40 0 −14) INI sensitivity negatively correlated with RTI performance INI sensitivity. (d) Plotted values of the peak voxel from the cluster in c. (e) Left Insula (MNI coordinates: −38 0 12) INI sensitivity positively corelated with Pattern recognition memory INI sensitivity. (f) Plotted values of the peak voxel from the cluster in e.

Next, we tested for associations with cognitive data collected on the scan days. Specifically, we regressed difference scores (insulin scores minus placebo scores) for each task showing a significant effect of group against the contrast of activation from insulin versus placebo. We identified a negative association between reaction time index and sensitivity of the left anterior superior temporal gyrus to insulin (MNI coordinates: −40, 0, 14; k=72; p_(FWE)_ =0.014) (Fig. 4c) such that greater increase in activity under insulin compared to placebo was associated with faster reaction time in insulin vs. placebo. A positive association was observed for pattern recognition memory and the sensitivity of the left anterior dorsal insula to insulin (MNI coordinates: −38, 0, 12; k=59; p_(FWE)_ =0.025) (Fig. 4e) such that greater increase in activity under insulin compared to placebo was associated with better pattern recognition memory in insulin vs. placebo. These results show that central insulin sensitivity is associated with faster reaction time (left anterior superior temporal gyrus) and better pattern recognition memory (left anterior dorsal insula).

#### Caudate connectivity and cognitive performance

Finally, to test our hypothesis that central insulin sensitivity may be related to DA function and impact DA-dependent cognition, we performed connectivity analyses using the caudate as a region of interest. Spatial working memory score from the neuropsychological testing day was regressed against caudate connectivity in insulin versus placebo (n = 28). A negative correlation between number of errors and caudate connectivity was observed in the right intraparietal sulcus (MNI coordinates: 38, −56, 58; k=127; p_(FWE)_ =0.018) (Fig. 5a and b) and bilateral precuneus (MNI coordinates: 6, −62, 44; k=114; p_(FWE)_ =0.03) (Fig. 5a and c). We also observed a positive correlation between performance on the VSPLOT task and caudate connectivity (n=37) with four clusters in the occipital lobe, including the right cuneus and left lingual gyrus (MNI coordinates: 8, −92, 26; k=521; p_(FWE)_ < 0.001; 12, −74, 2; k=340; p_(FWE)_<0.001; −16, −96, 30; k=101; p_(FWE)_ = 0.035; −12, −94, −10; k=124; p_(FWE)_ =0.011; Fig. 5d-e). This shows that performance on the visuospatial task is related to connectivity between the caudate and occipital as well as frontal lobes.

**Figure 5.**
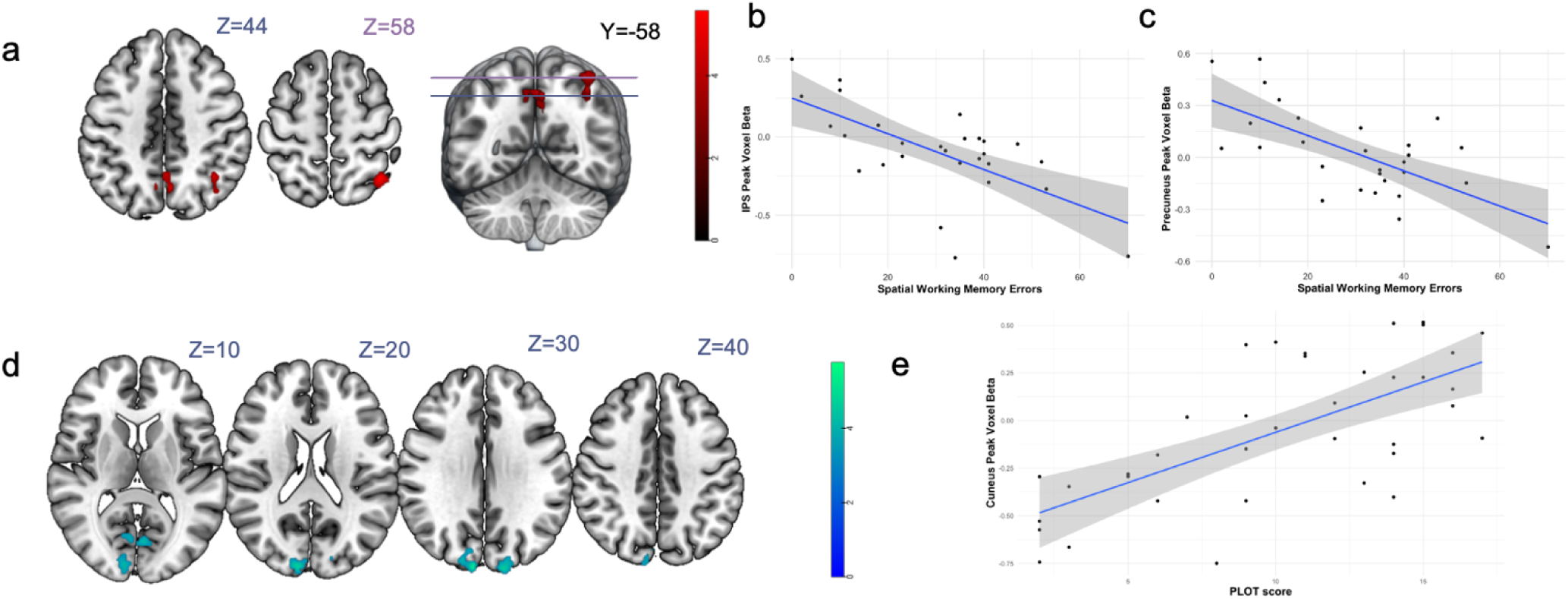
Caudate Connectivity sensitivity to INI associated with Spatial Working Memory and visuospatial processing. (a) Right intraparietal sulcus (MNI coordinates: 38 −56 58) and bilateral precuneus (MNI coordinates: 6 −62 44) connectivity with the caudate for insulin minus placebo shows a negative correlation with errors made on SWM task. (b) Plotted values of the peak voxel from the IPS cluster in A from the Insulin – Placebo map against SWM errors. (c) Plotted values of the peak voxel from the bilateral caudate cluster in a. (d) Right cuneus (MNI coordinates: 8, −92, 26) and left lingual gyrus (MNI coordinates: −12, −94, −10) connectivity with the caudate for insulin minus placebo shows a positive correlation with visuospatial processing. (e) Plotted values of the peak voxel from the right cuneus cluster against Penn Line Orientation Task (PLOT) score.

#### Cross-sectional brain structure

To identify structural brain differences between preT2D- and preT2D+ groups we used linear regression analyses with group, BMI z-scores, and Matsuda index as predictors of interest (separate models) and age, sex, and BMI z-scores (only for Matsuda index and group analyses) as covariates. At baseline, total surface area trended towards being larger in preT2D- (n=39, mean = 173949 ± 15905 mm^2^) compared to preT2D+ (n=20, mean = 163815 ± 13350 mm^2^), but this effect did not survive multiple comparisons correction (mean difference = −8,205 ± 3338 mm^2^, t(54) = −2.46, p = 0.0172, p = 0.0516). BMI z-score and Matsuda index were not related to anatomical measures (Supplement Table 4).

### Longitudinal results

#### Longitudinal characteristics of groups as defined using baseline classification

There were no differences in measures of demographics, diet, sleep, or depression at baseline or follow-up. Adiposity measures did not differ between groups at baseline, and only one measure differed at follow-up. Specifically, deep-to-superficial subcutaneous fat ratio was lower for preT2D+ (mean = 1.41 ± 0.29) compared to preT2D- (preT2D- mean = 1.76 ± 0.51) at Follow-up (p = 0.018), but not at Baseline (p = 0.054). Insulin sensitivity, as measured by Matsuda index, was lower for preT2D + (mean = 1.19 ± 0.46) compared to preT2D- (mean = 1.71 ± 1.15) at Baseline (p = 0.049), but not at Follow-up (p = 0.8). Full statistics are presented in Table 1.

#### Change in prediabetes status and cognitive variability over time

Some participants’ metabolic health improved over two years while others worsened and we hypothesized that these changes would be accompanied by greater variability in cognition (in either direction), as opposed to individuals whose metabolic health remained stable. To test this, we grouped participants who changed their metabolic health (either improved or worsened) into one bin, and those who did not change their metabolic health into another bin, calculated absolute baseline-to-follow-up cognitive score differences for each individual, and used linear regression analyses with relevant covariates to assess differences between the bins. There were no differences in the magnitude of cognitive change between individuals who with variable versus stable metabolic functions. See Supplement Table 5 for full result.

#### Effects of prediabetes status (as measured at baseline), insulin resistance, and adiposity on cognition over time

To investigate whether metabolic health was associated with changes in cognition over the 2-year follow-up period, we used linear mixed model regressions. The analysis revealed consistent impairments in general cognition and visuospatial processing for those with prediabetes at baseline. In general cognition, preT2D+ scored worse than preT2D- on WISC/WAIS composite scores for Full Scale IQ (Fig. 6a, F(1, 36.06) = 5.62, p = 0.0233, p_FDRadj_ = 0.035) and Fluid Reasoning Index (Fig. 6b, F(1, 35.70) = 11.98, p = 0.0014, p_FDRadj_ = 0.0042). Similarly, for visuospatial reasoning, Block Design score from the WISC/WAIS assessment was lower in preT2D+ than preT2D- (Fig. 6c, F(1, 36.51) = 7.65, p = 0.0089, p_FDRadj_ = 0.0177), and preT2D+ had more errors on VSPLOT than preT2D- (Fig. 6d, F(1, 34.70) = 6.01, p = 0.0194, p_FDRadj_ = 0.0194).

**Figure 6.**
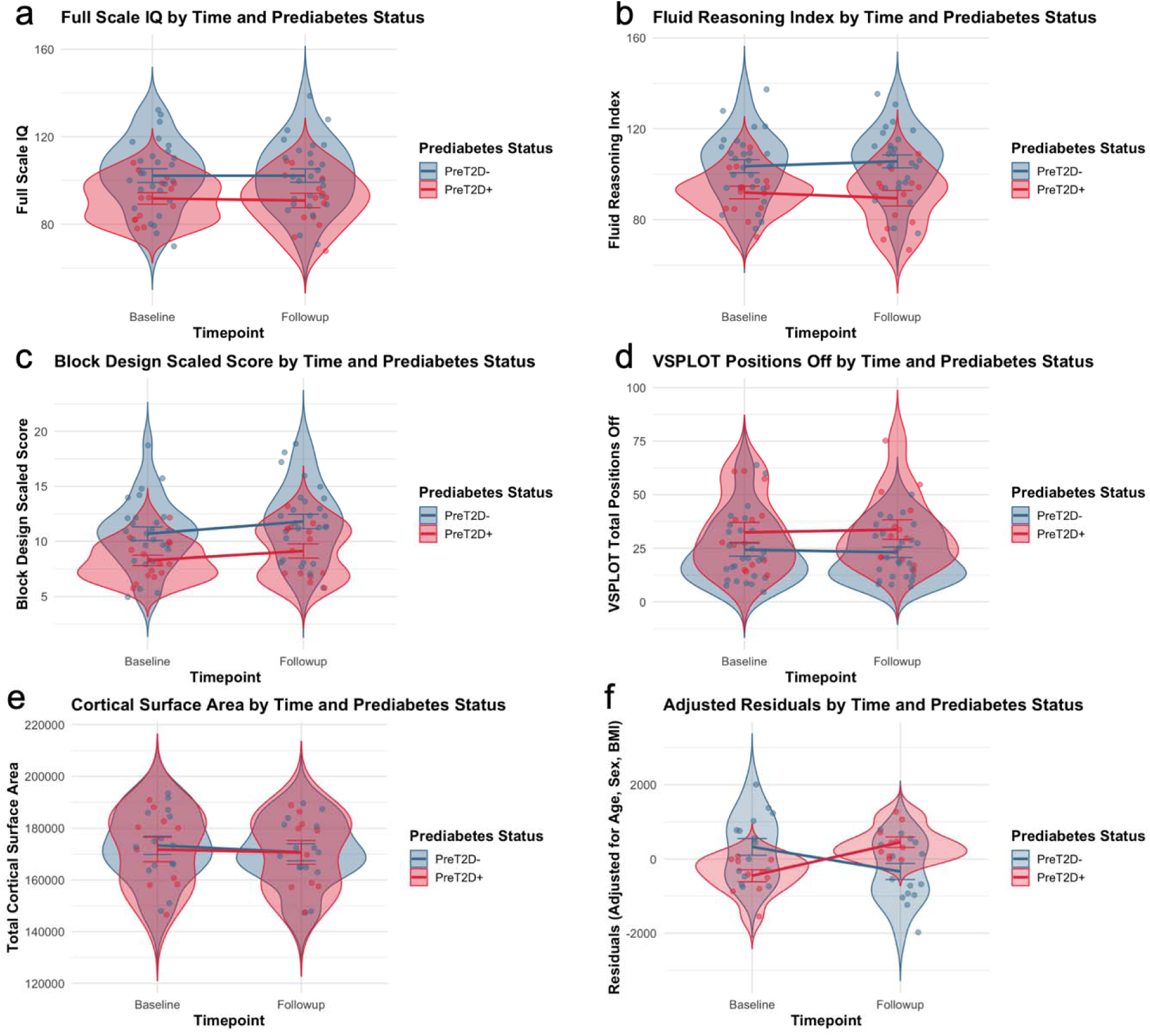
Longitudinal cognitive function and brain structure in relation to baseline prediabetes status. In mixed effects linear models, participants with prediabetes at baseline showed stable impairments in general cognition as measured by Full Scale IQ (a) and Fluid Reasoning Index (b) scores from the WISC/WAIS assessments. Similarly, this group consistently performed worse on the WISC/WAIS Block Design task (c) and the Variable Short Penn Line Orientation Test (d). There were no interactions between group and time for any of these tasks. Mixed effect linear regressions revealed a significant prediabetes group X time interaction for total cortical surface area (e). Residuals after adjusting for age, sex, and BMI as covariates in this model show this interaction indicates a greater decrease in surface area for the no prediabetes group (f).

There was a trending BMI by time interaction for Digit Span score (F(1, 40.25) = 5.51, p = 0.0239, p_FDRadj_ = 0.0956) and a trending Matsuda index main effect on Trail Making Test A score (F(1, 70.31) = 5.10, p = 0.027, p_FDRadj_ = 0.081), but neither survived multiple comparisons correction. The sample did not show any other associations between cognition and BMI or Matsuda index for main effects or interactions with time (Supplement Tables 6-8).

#### Longitudinal Brain Structure

We also tested whether there are structural brain changes over the 2-year follow-up period by examining longitudinal differences in the variables of interest. Linear mixed model regressions revealed a significant group by time interaction on total surface area (Fig. 6e-f, F(1, 22.35) = 6.93, p = 0.0151, p_FDRadj_ = 0.0453), where participants who were originally grouped as preT2D-had a greater decrease in surface area at follow-up compared to those grouped as preT2D+ (Supplement Table 9).

Surface area was not correlated with IQ at baseline or longitudinally, and there were no interactions with time. Including IQ as a covariate in models investigating the relationship between group and surface area marginally increased the significance of effects of group at baseline (mean difference = −8,922.0 ± 3355.5 mm^2^, t(53) = −2.659, p = 0.0103) and group by time interaction in longitudinal analysis (F(1, 21.54) = 7.09, p = 0.0143). No other effects were found for the group, BMI, or Matsuda index models on the anatomical measures assessed.

## Discussion

In this longitudinal study of youth with overweight and obesity, we demonstrate that prediabetes is associated with broad differences in cognitive function that are independent of adiposity and remain stable over a two-year period. These findings provide evidence that the neurocognitive differences linked to metabolic dysfunction are present early in development and persist across time.

At baseline, youth with prediabetes exhibited lower global cognitive performance as well as deficits in executive function, psychomotor speed, and visuospatial processing. These differences were observed after controlling for adiposity, suggesting that metabolic dysfunction contributes to neurocognitive variation beyond the effects of obesity alone. Importantly, when accounting for global cognitive ability, most domain-specific effects were attenuated, with the exception of spatial working memory. This pattern suggests that prediabetes may be associated with a broad shift in cognitive performance rather than highly selective deficits, consistent with a general influence on neurocognitive function during development. In addition, across the full sample, reduced peripheral insulin sensitivity was associated with slower processing speed, suggesting a relationship between metabolic function and cognitive performance that extends beyond categorical group differences.

While prior work in adults with T2D has consistently reported impairments in similar domains, it has remained unclear whether these deficits emerge only after prolonged disease exposure or reflect earlier developmental processes [9], [10], [11], [23]. Our findings extend this literature by showing that the cognitive differences are already present at the stage of prediabetes in youth even after controlling for adiposity. This suggests that the poorer cognitive performance associated with prediabetes may exist prior to, or develop very early in the onset of the course of the disease.

A key contribution of this study is the longitudinal demonstration that cognitive differences between youth with and without prediabetes are stable over a two-year period. These differences were not explained by changes in adiposity or glucose tolerance, and were observed despite variability in metabolic trajectories across individuals. This stability argues against a model in which short-term fluctuations in metabolic status directly drive corresponding changes in cognition. This interpretation is further supported by the structural imaging results. While youth with normal glucose control at baseline exhibited the expected developmental reductions in cortical surface area [44], this pattern was not observed in youth with prediabetes. Although these findings should be interpreted cautiously, they raise the possibility that prediabetes is associated with alterations in typical neurodevelopmental trajectories.

Functional imaging analyses were limited by attrition and motion-related data loss; however, exploratory analyses collapsing across groups revealed several convergent findings. Peripheral insulin sensitivity was positively associated with the neural response to intranasal insulin in the intraparietal sulcus, independent of adiposity and demographic covariates. Insulin-related activity in this region was also linked to cognitive performance, such that stronger insulin-associated connectivity between the intraparietal sulcus and caudate was associated with better working memory. We also observed a similar effect with caudate connectivity to the precuneus, a region previous shown to have dose dependent responses to INI in adults [45].

Given evidence that caudate responses to intranasal insulin are associated with dopamine signaling [14], these findings are broadly consistent with the hypothesis that insulin action on dopaminergic circuits may link metabolic dysfunction to cognitive performance [46]. Related frameworks have also been proposed in other domains, including depression, where insulin–dopamine interactions have been implicated in pathophysiology [47]. However, given the limited sample size and exploratory nature of these analyses, these findings should be interpreted cautiously.

Overall, our functional imaging results are consistent with prior work in adults showing that better peripheral glucose control is associated with greater brain sensitivity to intranasal insulin [48], [49], [50] and extend this literature by implicating insulin action on corticostriatal circuits with cognitive function.

### Limitations

Our study has several limitations. Most critically, our sample size was relatively small due to the impact of the COVID-19 pandemic on recruitment and retention. In addition, a number of participants obtained braces following the baseline assessment, rendering them ineligible for the follow-up neuroimaging measurements. The resulting decrease in sensitivity raises the possibility that subtle effects between groups or timepoints were present but not detectable.

Furthermore, our ancillary study design did not contain a healthy-weight control group. Therefore, the effects that we did observe may have resulted from an interaction between adiposity and poor glucose control. Finally, we were approved to administer 1 IU per kg up to a maximum dose of 60 UI. However, based on body weight, all participants met criteria for the highest dose, which might have been too low to reach efficacy; especially for the participants with higher body weight.

### Conclusions and future directions

Together, these findings demonstrate that prediabetes in youth with overweight and obesity is associated with broad and persistent differences in cognitive function that are independent of adiposity and stable over time. The early emergence and stability of these differences suggest that neurocognitive alterations may be a feature of metabolic risk prior to the onset of overt disease. Consistent with this interpretation, structural imaging results indicate a deviation from expected developmental reductions in cortical surface area in youth with prediabetes, raising the possibility of altered neurodevelopmental trajectories. While exploratory functional neuroimaging results link peripheral insulin resistance with central insulin sensitivity in youth and point to a potential role for insulin sensitivity of corticostriatal circuitry in working memory function.

## Data Availability

De-identified data may be accessed on a case-by-case basis by contacting the corresponding author.

## Acknowledgements

We would like to thank all participants and project members for their participation in this study.

## Data Availability

De-identified data may be accessed on a case-by-case basis by contacting the corresponding author.

## Author’s relationships and activities

All authors declare that they have no relationships or activities that might bias, or be perceived to bias, their work on this study.

## Contribution statement

SC and DMS were involved in the conception and design of the study. JQ, TA, JS, TK, XD, NS, BP, SC, and DS were involved in the conduct of the study and acquisition of data. JQ, FM, AC, KM, FL, SK, HP NS, AD, SC, and DS were involved in the analysis and interpretation of results. JQ, FM, and DMS wrote the first draft of the manuscript with rsFMRI Methods written by AC. All authors edited, reviewed, and approved the final version of the manuscript.

## Supplementary Figures

**Supplement Figure 1.**
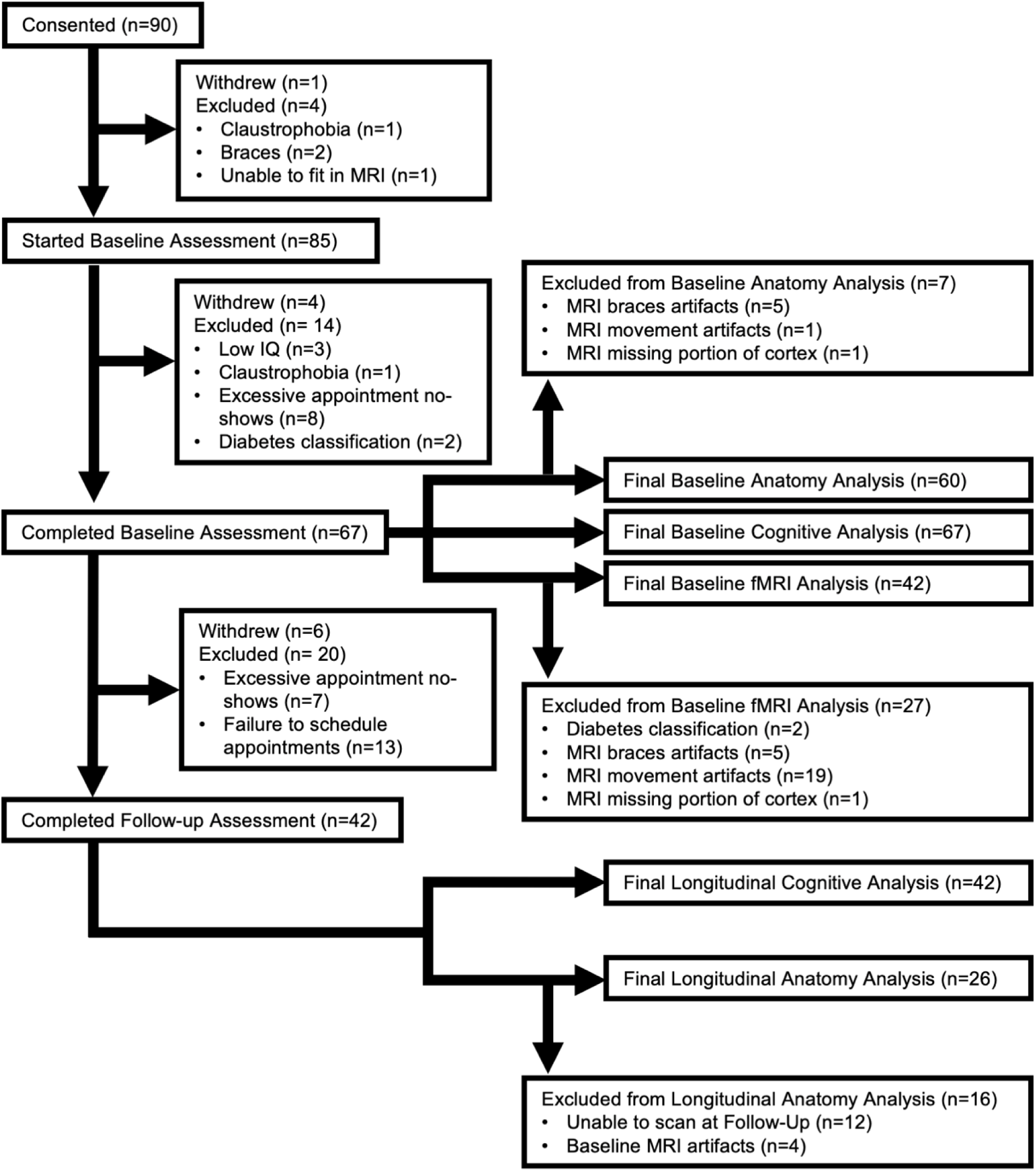
Participant flow diagram. Participant numbers showing the withdrawals and exclusions from the study from the consenting process, through the Follow-Up Assessment, to the analyses.

**Supplement Figure 2.**
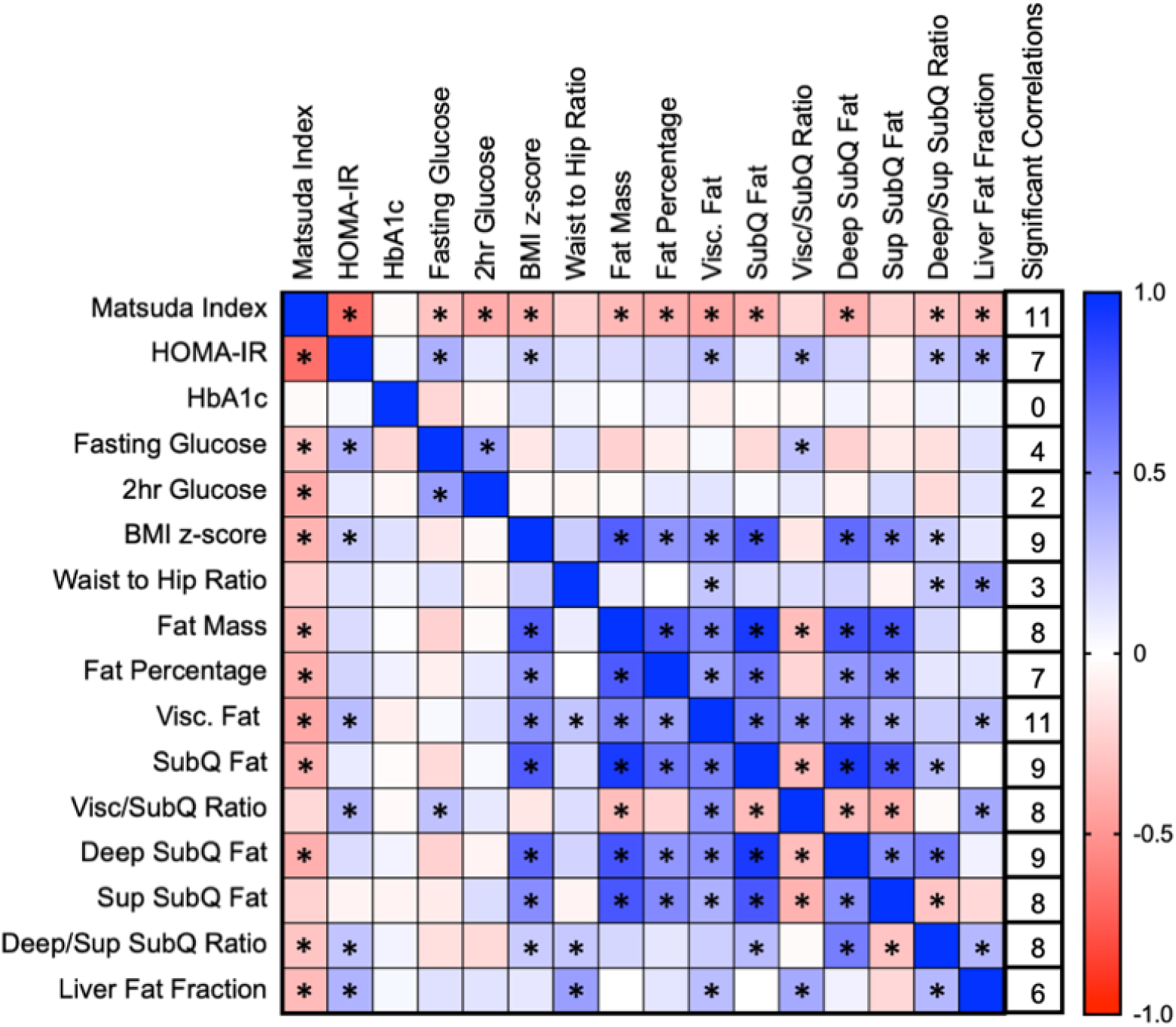
Correlations matrix of metabolic and adiposity measures. Significant Pearson correlations (p<0.05) are marked with an “*”. Box color indicates strength and direction of correlation where blue colored boxes indicate a positive relationship between variables and red colored boxes indicate a negative relationship. Abbreviations: Visceral Fat – Visc. Fat; Subcutaneous Fat – SubQ Fat; Visceral fat to subcutaneous fat ratio – Visc/SubQ Ratio; Deep subcutaneous fat – Deep SubQ Fat; Superficial subcutaneous fat – Sup SubQ Fat; Deep subcutaneous fat to superficial subcutaneous fat ratio – Deep/Sup SubQ Ratio.

## Supplementary Tables

**Supplement Table 1.** Additional baseline measurements. : Total Household Income, HOMA-IR, HbA1c, Fasting Glucose, Glucose at 120min, CES Depression Scale for Children, and Berlin Sleep Apnea Questionnaire

| Characteristic | Whole Cohort, N = 66 | preT2D-, N = 44 | preT2d+, N = 22 | *p*-value |
| --- | --- | --- | --- | --- |
| Total Household Income |  |  |  | 0.27<sup>3</sup> |
| <\$25,000 | 12 (25) | 9 (28) | 3 (19) | |
| \$25,000-\$50,000 | 15 (31) | 11 (34) | 4 (25) | |
| \$50,000-\$75,000 | 10 (21) | 6 (19) | 4 (25) | |
| \$75,000-\$100,000 | 4 (8.3) | 1 (3.1) | 3 (19) | |
| \$100,000-\$150,000 | 6 (13) | 5 (16) | 1 (6.3) | |
| >\$150,0000 | 1 (2.1) | 0 (0) | 1 (6.3) | |
| Declined to respond | 18 | 12 | 6 |  |
| CES Depression Scale for Children | 17 (12) | 18 (12) | 15 (12) | 0.301 |
| missing | 3 | 1 | 2 |  |
| Berlin Sleep Apnea Questionnaire | 1.36 (0.48) | 1.33 (0.48) | 1.4 (0.50) | 0.621 |
| missing | 7 | 5 | 2 |  |
| 1 Welch Two Sample t-test |  |  |  |  |
| 2 Pearson's Chi-squared test |  |  |  |  |
| 3 Fisher's exact test |  |  |  |  |
| Data are n (%) or mean ± SD unless otherwise noted. |  |  |  |  |

**Supplement Table 2.** Cross-sectional group differences in cognition. Linear regressions were run to assess cognition in adolescents classified as having prediabetes (preT2D+) or not (preT2D-) at baseline.

|  |  | preT2D- |  | preT2D+ |  | Model 1 |  | Model 2 (IQ included as covariate) |  |
| --- | --- | --- | --- | --- | --- | --- | --- | --- | --- |
| Domain | Task | n | Mean ± SD | n | Mean ± SD | Estimate ± SE | p-value | Estimate ± SE | p-value |
| Composite Cognition Scores | Verbal Comprehension Index (WISC/WAIS) | 44 | 99.36 ± 17.22 | 22 | 95.14 ± 9.63 | -4.045 ± 3.991 | 0.315 | 2.015 ± 2.552 | 0.433 |
| | Fluid Reasoning Index (WISC/WAIS) | 44 | 100.82 ± 14.69 | 22 | 94.05 ± 11.00 | -6.848 ± 3.633 | 0.0642 | -0.669 ± 1.773 | 0.707 |
| Executive Function | Spatial Working Memory (CANTAB) | 29 | 24.66 ± 13.21 | 16 | 40.00 ± 12.61 | 14.753 ± 4.036 | **<0.001** | 12.734 ± 4.131 | **0.00376** |
| | Stop Signal Task (CANTAB) | 41 | 254.36 ± 65.79 | 21 | 298.01 ± 85.71 | 44.300 ± 18.133 | **0.0177** | 35.363 ± 18.287 | 0.0582 |
| | Digit Span (WISC/WAIS) | 44 | 9.52 ± 2.31 | 22 | 8.41 ± 2.89 | -1.115 ± 0.667 | 0.0997 | -0.344 ± 0.554 | 0.537 |
| | Trail Making Test B | 44 | 60.65 ± 27.10 | 21 | 69.60 ± 28.89 | 9.244 ± 7.101 | 0.198 | 4.913 ± 6.858 | 0.477 |
| Temporal Discounting | Delayed Discounting | 33 | -4.40 ± 2.16 | 20 | -3.63 ± 1.97 | 0.774 ± 0.590 | 0.196 | 0.340 ± 0.560 | 0.547 |
| Psychomotor Speed | Reaction Time Index (CANTAB) | 42 | 251.81 ± 56.05 | 21 | 291.57 ± 56.49 | 39.909 ± 14.903 | **0.00962** | 27.227 ± 14.007 | 0.0569 |
| | Trail Making Test A | 44 | 24.33 ± 9.90 | 21 | 28.02 ± 10.02 | 3.831 ± 2.430 | 0.12 | 2.611 ± 2.401 | 0.281 |
| | Coding (WISC/WAIS) | 44 | 9.64 ± 3.41 | 22 | 9.18 ± 3.33 | -0.541 ± 0.837 | 0.52 | 0.466 ± 0.678 | 0.495 |
| Memory | Pattern Recognition Memory (CANTAB) | 26 | 77.14 ± 17.80 | 15 | 65.92 ± 18.05 | -9.826 ± 5.604 | 0.088 | -6.845 ± 5.678 | 0.236 |
|  | Delayed Matching to Sample (CANTAB) | 42 | 79.00 ± 14.80 | 21 | 74.33 ± 16.82 | -4.918 ± 3.380 | 0.151 | -3.725 ± 3.465 | 0.287 |
| Visuospatial Processing | Block Design (WISC/WAIS) | 44 | 10.09 ± 3.05 | 22 | 8.41 ± 1.79 | -1.672 ± 0.723 | **0.0241** | -0.766 ± 0.570 | 0.184 |
|  | Variable Short Penn Line Orientation Test | 41 | 25.34 ± 14.57 | 21 | 33.62 ± 17.03 | 8.647 ± 3.843 | **0.0283** | 5.934 ± 3.532 | 0.0985 |

**Supplement Table 3.**
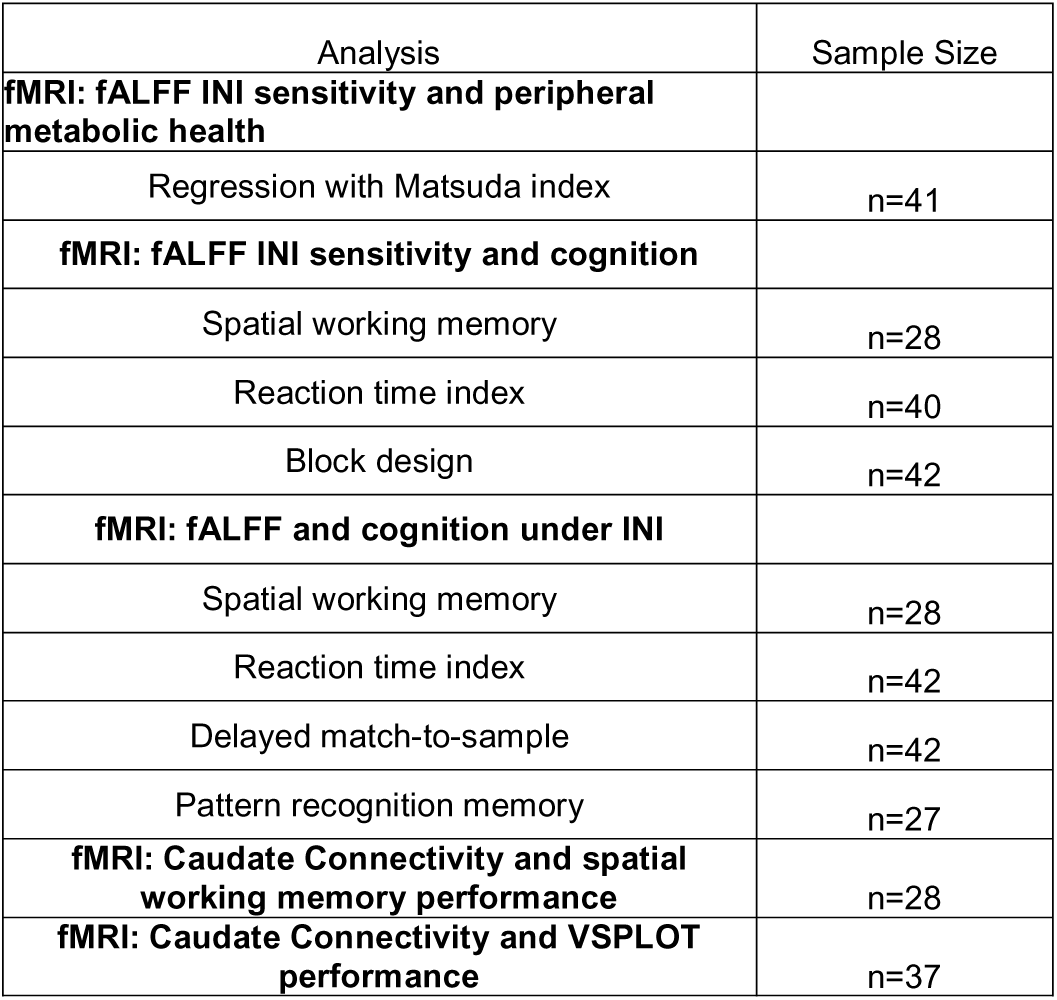
Sample size summary for fMRI analyses

**Supplement Table 4.** Effects of baseline prediabetes status on brain structure at baseline

| Outcome | preT2D- n=39<br>Mean ± SD | preT2D+ n=20<br>Mean ± SD | Estimate | SE | *p*-value |
| --- | --- | --- | --- | --- | --- |
| Mean cortical thickness | 2.65 ± 0.08 | 2.65 ± 0.09 | 0.00 | 0.02 | 0.995 |
| Cortical surface area | 173948.95 ± 15905.54 | 163815.12 ± 13350.63 | -8204.99 | 3338.50 | 0.0172 |
| IPS cortical thickness | 2.37 ± 0.1 | 2.38 ± 0.16 | 0.03 | 0.03 | 0.399 |
| Hippocampal Volume | 0.01 ± 0 | 0.01 ± 0 | 0.00 | 0.00 | 0.179 |

**Supplement Table 5.**
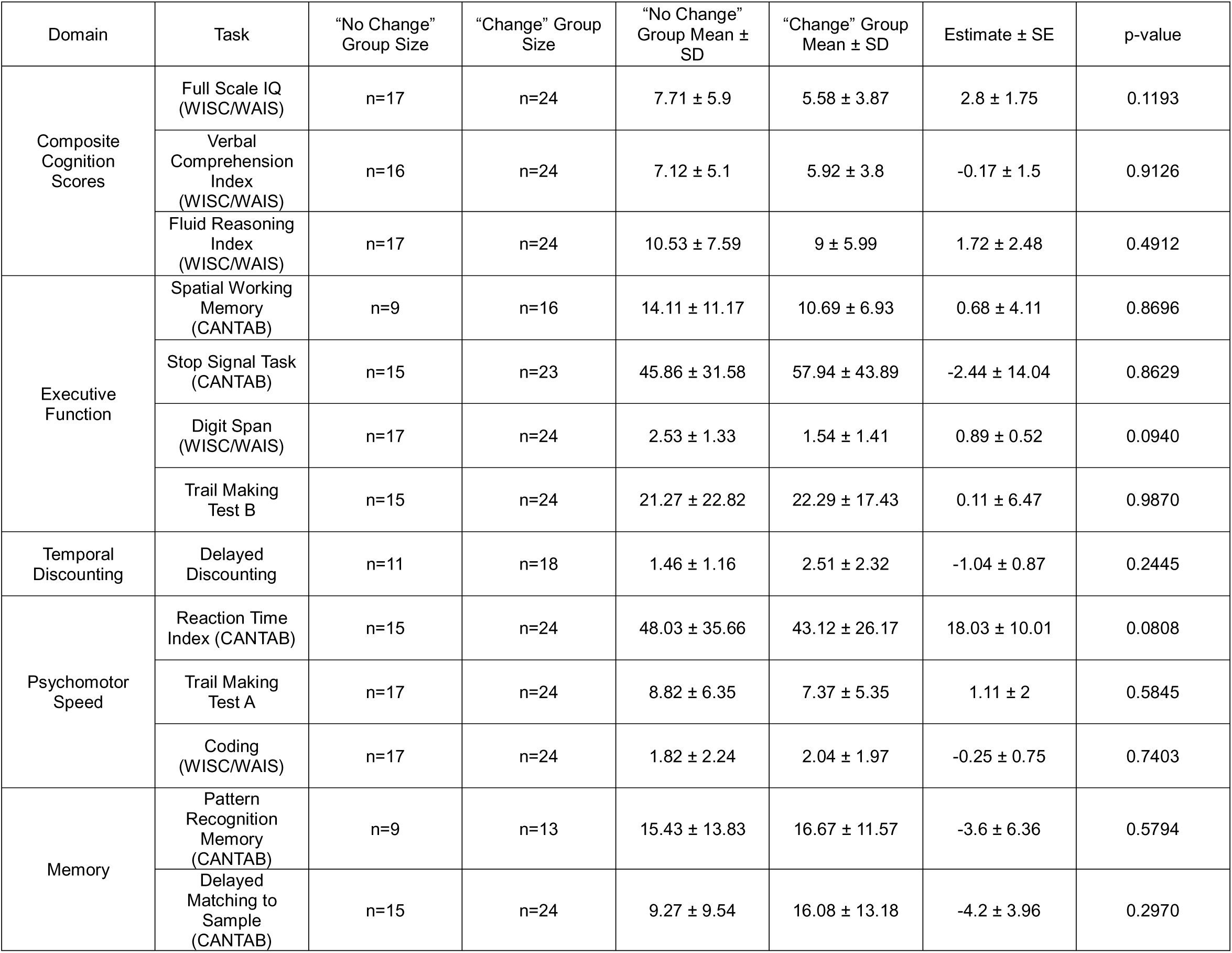
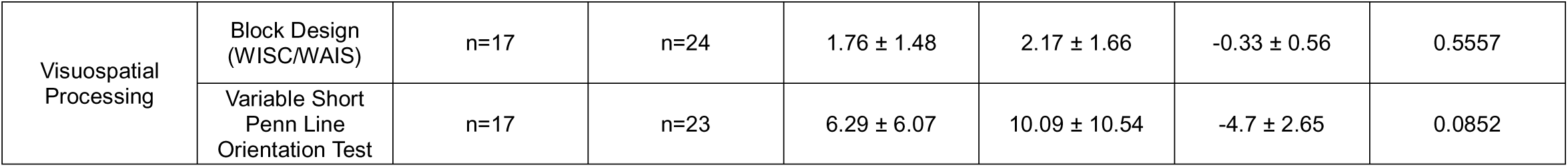
Change in prediabetes status and cognitive variability over time. Comparison of absolute differences in cognition for groups who changed metabolic status (“Change” group) or maintained metabolic status (“No Change” group) over two years.

**Supplement Table 6.** Effects of baseline prediabetes status on cognition. Cognition in adolescents classified as having prediabetes (preT2D+) or not (preT2D-) at baseline. Linear mixed effects models were run to assess effects of baseline group classification on cognitive performance (group effect), as well as change in cognition over time (group X time interaction). Bold values depict p-values surviving multiple comparisons correction.

| Domain | Task | preT2D- Baseline |  | preT2D- Follow-up |  | preT2D+ Baseline |  | preT2D+ Follow-up |  | Group Effect |  | Group X Time Interaction |  |
| --- | --- | --- | --- | --- | --- | --- | --- | --- | --- | --- | --- | --- | --- |
| | | n | Mean ± SD | n | Mean ± SD | n | Mean ± SD | n | Mean ± SD | F | p-value | F | p-value |
| Composite Cognition Scores | Full Scale IQ (WISC/WAIS) | 27 | 102.19 ± 16.34 | 27 | 102.19 ± 15.97 | 14 | 91.79 ± 9.86 | 14 | 90.86 ± 12.21 | 5.62 | **0.0233** | 0.32 | 0.5744 |
| | Verbal Comprehension Index (WISC/WAIS) | 26 | 99.42 ± 16.67 | 27 | 100.63 ± 17.39 | 14 | 94.36 ± 7.63 | 14 | 93 ± 10.65 | 2.46 | 0.126 | 0.59 | 0.446 |
| | Fluid Reasoning Index (WISC/WAIS) | 27 | 103.48 ± 15 | 27 | 105.63 ± 14.77 | 14 | 91.93 ± 10.36 | 14 | 89.43 ± 12.71 | 11.98 | **0.00141** | 1.53 | 0.22293 |
| Executive Function | Spatial Working Memory (CANTAB) | 17 | 28.82 ± 12.13 | 27 | 22.63 ± 13.02 | 8 | 42.62 ± 7.54 | 14 | 25.14 ± 11.27 | 4.03 | 0.0521 | 3.22 | 0.0836 |
| | Stop Signal Task (CANTAB) | 25 | 251.34 ± 68.59 | 27 | 249.46 ± 60 | 13 | 275.63 ± 92.84 | 14 | 248.52 ± 89.11 | 0.78 | 0.3840 | 1.55 | 0.2211 |
| | Digit Span (WISC/WAIS) | 27 | 9.63 ± 2.68 | 27 | 9.52 ± 2.06 | 14 | 8 ± 2.29 | 14 | 8.5 ± 2.38 | 3.40 | 0.0736 | 0.66 | 0.4214 |
| | Trail Making Test B | 26 | 57.2 ± 26.5 | 27 | 52.67 ± 16.72 | 13 | 68.02 ± 24.21 | 13 | 59.05 ± 24.04 | 2.14 | 0.1532 | 0.09 | 0.7658 |
| Temporal Discounting | Delayed Discounting | 20 | -4.31 ± 2.1 | 23 | -4.88 ± 2.24 | 13 | -3.42 ± 1.96 | 11 | -5.36 ± 1.73 | 0.06 | 0.8075 | 1.90 | 0.1782 |
| Psychomotor Speed | Reaction Time Index (CANTAB) | 26 | 254.1 ± 58.41 | 27 | 236.78 ± 52.06 | 13 | 284.85 ± 53.16 | 14 | 261.71 ± 63.36 | 2.59 | 0.116 | 0.12 | 0.736 |
| | Trail Making Test A | 27 | 24.02 ± 10.95 | 27 | 24.79 ± 9.08 | 14 | 26.23 ± 8.38 | 14 | 23.72 ± 10.26 | 0.00 | 0.9993 | 0.97 | 0.3296 |
|  | Coding<br>(WISC/WAIS) | 27 | 10.22 ± 3.81 | 27 | 9.37 ± 3.52 | 14 | 10 ± 3.37 | 14 | 9.36 ± 3.82 | 0.00 | 0.948 | 0.00 | 0.961 |
| Memory | Pattern<br>Recognition<br>Memory<br>(CANTAB) | 15 | 75.56 ± 18.04 | 27 | 77.57 ± 19.34 | 8 | 67.36 ± 19.34 | 13 | 65.81 ± 18.82 | 3.07 | 0.0885 | 0.47 | 0.4986 |
|  | Delayed<br>Matching to<br>Sample<br>(CANTAB) | 26 | 76.85 ± 15.44 | 27 | 79.52 ± 18.22 | 13 | 77.46 ± 15.59 | 14 | 71 ± 22.53 | 1.77 | 0.191093 | 2.02 | 0.163123 |
| Visuospatial<br>Processing | Block Design<br>(WISC/WAIS) | 27 | 10.7 ± 3.21 | 27 | 11.81 ± 3.39 | 14 | 8.29 ± 1.77 | 14 | 9.14 ± 2.41 | 7.65 | **0.00887** | 0.12 | 0.72910 |
|  | Variable Short<br>Penn Line<br>Orientation Test | 27 | 24.33 ± 15.54 | 26 | 23.15 ± 12.44 | 14 | 32.43 ± 17.35 | 14 | 33.71 ± 17.1 | 6.01 | **0.01936** | 0.17 | 0.67965 |

**Supplement Table 7.** Effects of BMI on cognition over time. Linear mixed effects models were run to assess effects of BMI on cognitive performance, as well as change in cognition over time (BMI X time interaction). Bold values depict p-values surviving multiple comparisons correction.

| Variable | Baseline n | Baseline Mean ± SD | Follow-up n | Follow-up Mean ± SD | BMI Effect F | BMI Effect *p*-value | BMI X Time Interaction F | BMI X Time Interaction *p*-value |
| --- | --- | --- | --- | --- | --- | --- | --- | --- |
| Full Scale IQ (WISC/WAIS) | 41 | 98.63 ± 15.17 | 41 | 98.32 ± 15.62 | 0.52 | 0.473 | 1.76 | 0.193 |
| Verbal Comprehension Index (WISC/WAIS) | 40 | 97.65 ± 14.27 | 41 | 98.02 ± 15.71 | 0.50 | 0.483 | 0.05 | 0.823 |
| Fluid Reasoning Index (WISC/WAIS) | 41 | 99.54 ± 14.56 | 41 | 100.1 ± 15.96 | 0.00 | 0.973 | 0.15 | 0.702 |
| Spatial Working Memory (CANTAB) | 25 | 33.24 ± 12.56 | 41 | 23.49 ± 12.36 | 0.52 | 0.475 | 0.27 | 0.609 |
| Stop Signal Task (CANTAB) | 38 | 259.65 ± 77.35 | 41 | 249.14 ± 70.15 | 0.29 | 0.5952 | 0.72 | 0.3998 |
| Digit Span (WISC/WAIS) | 41 | 9.07 ± 2.64 | 41 | 9.17 ± 2.2 | 0.71 | 0.4025 | 5.51 | 0.0239 |
| Trail Making Test B | 39 | 60.81 ± 25.96 | 40 | 54.74 ± 19.32 | 2.13 | 0.1521 | 0.54 | 0.4688 |
| Delayed Discounting | 33 | -3.96 ± 2.06 | 34 | -5.03 ± 2.07 | 0.33 | 0.571 | 0.06 | 0.804 |
| Reaction Time Index (CANTAB) | 39 | 264.35 ± 57.9 | 41 | 245.29 ± 56.65 | 1.05 | 0.311 | 0.02 | 0.877 |
| Trail Making Test A | 41 | 24.77 ± 10.09 | 41 | 24.42 ± 9.39 | 0.05 | 0.8206 | 0.14 | 0.7122 |
| Coding (WISC/WAIS) | 41 | 10.15 ± 3.62 | 41 | 9.37 ± 3.58 | 1.38 | 0.245 | 0.03 | 0.862 |
| Pattern Recognition Memory (CANTAB) | 23 | 72.71 ± 18.49 | 40 | 73.75 ± 19.73 | 0.44 | 0.509 | 0.04 | 0.835 |
| Delayed Matching to Sample (CANTAB) | 39 | 77.05 ± 15.29 | 41 | 76.61 ± 19.93 | 0.04 | 0.839276 | 0.62 | 0.435449 |
| Block Design (WISC/WAIS) | 41 | 9.88 ± 3.01 | 41 | 10.9 ± 3.32 | 0.00 | 0.966 | 0.04 | 0.838 |
| Variable Short Penn Line Orientation Test | 41 | 27.1 ± 16.43 | 40 | 26.85 ± 14.92 | 0.15 | 0.69815 | 0.02 | 0.90302 |

**Supplement Table 8.**
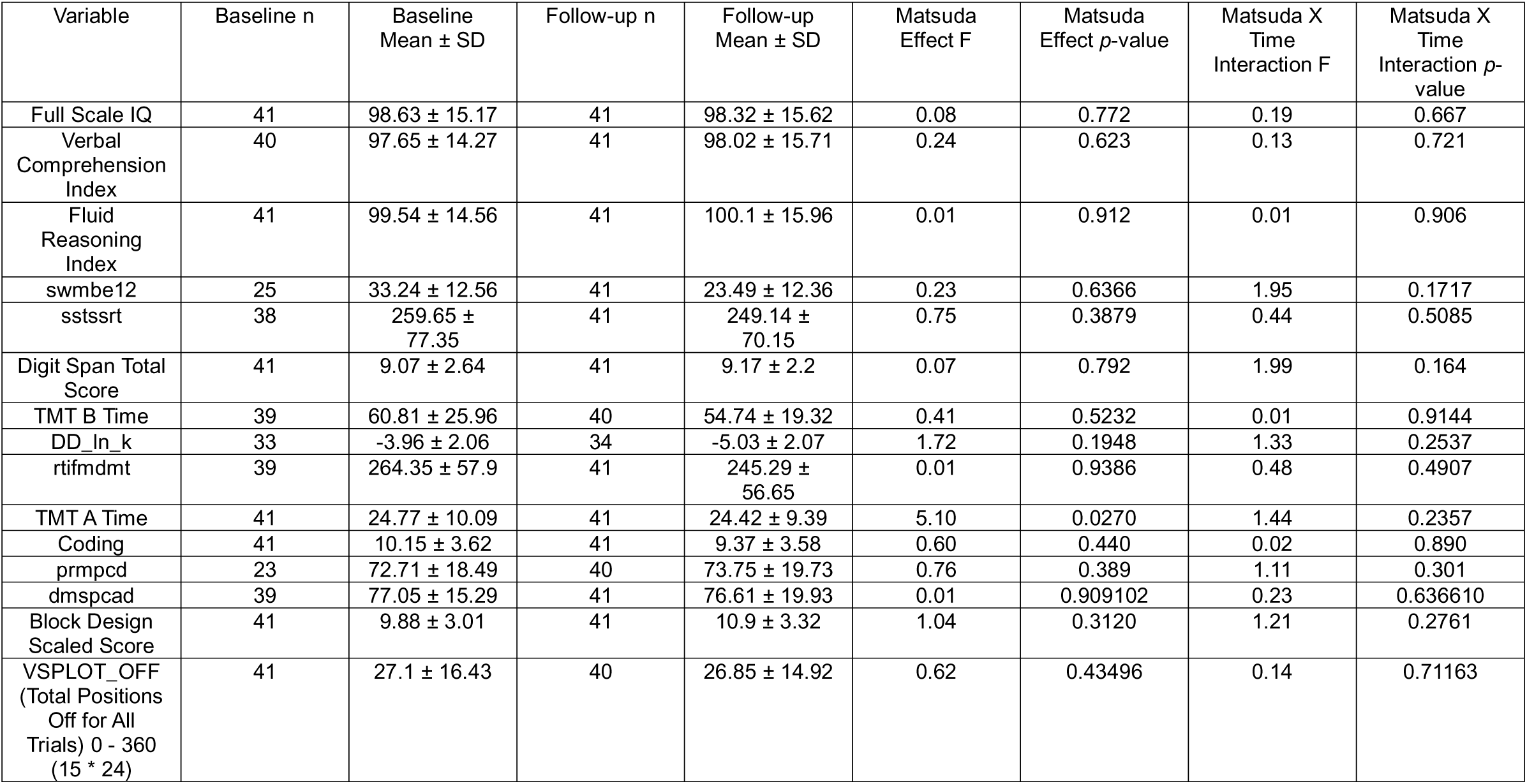
Effects of Matsuda Index on cognition over time. Linear mixed effects models were run to assess effects of Matsuda Index on cognitive performance, as well as change in cognition over time (Matsuda Index X time interaction). Bold values depict p-values surviving multiple comparisons correction.

**Supplement Table 9.** Effects of baseline prediabetes status on brain structure at baseline VS. 2-year follow-up.

| Outcome | preT2D- n=15 |  | preT2D+ n=10 |  | Group Effect |  | Group X Time Interaction |  |
| --- | --- | --- | --- | --- | --- | --- | --- | --- |
| | Baseline<br>Mean ± SD | Follow-up<br>Mean ± SD | Baseline<br>Mean ± SD | Follow-up<br>Mean ± SD | F | p-value | F | p-value |
| Mean cortical thickness | 2.676 ± 0.096 | 2.664 ± 0.075 | 2.694 ± 0.08 | 2.642 ± 0.084 | 0.01 | 0.925073 | 2.36 | 0.138103 |
| Cortical surface area | 173392.927 ± 13671.531 | 170701.593 ± 12700.437 | 171763.21 ± 14984.416 | 170660.99 ± 14483.629 | 0.02 | 0.8775 | 6.93 | **0.0151** |
| IPS cortical thickness | 2.397 ± 0.112 | 2.367 ± 0.088 | 2.407 ± 0.101 | 2.34 ± 0.085 | 0.01 | 0.93065 | 1.91 | 0.18046 |
| Hippocampal Volume | 0.006 ± 0 | 0.006 ± 0 | 0.006 ± 0.001 | 0.006 ± 0.001 | 0.90 | 0.353 | 0.37 | 0.549 |

